# Perivascular adipose tissue attenuation in CT angiography and plaque inflammation in patients with carotid atherosclerosis

**DOI:** 10.64898/2026.09.27.26364115

**Authors:** Stefan Mayrhofer, Luka Živković, Yingle Li, Roya Batool, Julian Louma, Paul Reidler, Abdalla Marei, Nikolaos Tsilimparis, DEMDAS study group, Marios K. Georgakis

## Abstract

**Background and Aims:** Quantifying arterial inflammation can augment risk stratification and drug development in atherosclerosis. While perivascular adipose tissue (PVAT) density, captured as fat attenuation index (FAI) on CT angiography (CTA), is an emerging biomarker of coronary inflammation, it remains unclear whether this concept extends to carotid atherosclerosis.

**Methods:** Using routine CTA data from hospital-based stroke cohorts, we developed a pipeline to quantify attenuation metrics of PVAT surrounding the distal common and proximal internal carotid arteries. We compared PVAT metrics between culprit and contralateral non-culprit plaques in patients with symptomatic carotid atherosclerosis and between culprit plaques and control carotid segments from patients without symptomatic atherosclerosis. In patients undergoing carotid endarterectomy, we assessed associations of CTA-derived PVAT metrics with histological and proteomic markers of plaque inflammation and stability.

**Results:** Across 107 patients with symptomatic carotid atherosclerosis (75.5±8.7 years; 63% men), PVAT attenuation metrics did not show significant differences between culprit and contralateral non-culprit lesions (all p>0.2). We also found no evidence for higher PVAT attenuation in culprit lesions, when compared to arteries of patients without symptomatic atherosclerosis. In 82 patients undergoing endarterectomy, pre-operative CTA-derived metrics of PVAT attenuation showed no significant correlations with CD68+ macrophage areas in excised plaques (r -0.11 to 0.07, all p>0.3). Similarly, no PVAT attenuation metrics were associated with thinner fibrous cap or higher plaque levels of pro-inflammatory cytokines (IL-6, TNF).

**Conclusions:** In contrast to coronary atherosclerosis, carotid PVAT attenuation was not associated with symptomatic culprit lesions or histological and molecular markers of plaque inflammation and vulnerability.

**Structured Graphical Abstract:** *Key Question:* Can perivascular adipose tissue (PVAT) attenuation on CT, proposed as a marker of coronary artery inflammation, also capture atherosclerotic plaque inflammation and vulnerability in the carotid arteries?

*Key Finding:* In 107 patients with carotid atherosclerosis-associated stroke, PVAT attenuation did not differ between culprit and contralateral non-culprit lesions and was not associated with histopathological or molecular markers of plaque inflammation and vulnerability.

*Take-home Message:* PVAT attenuation did not capture local carotid plaque inflammation or vulnerability in this cohort, questioning the transferability of this CT-based marker from coronary to carotid atherosclerosis. 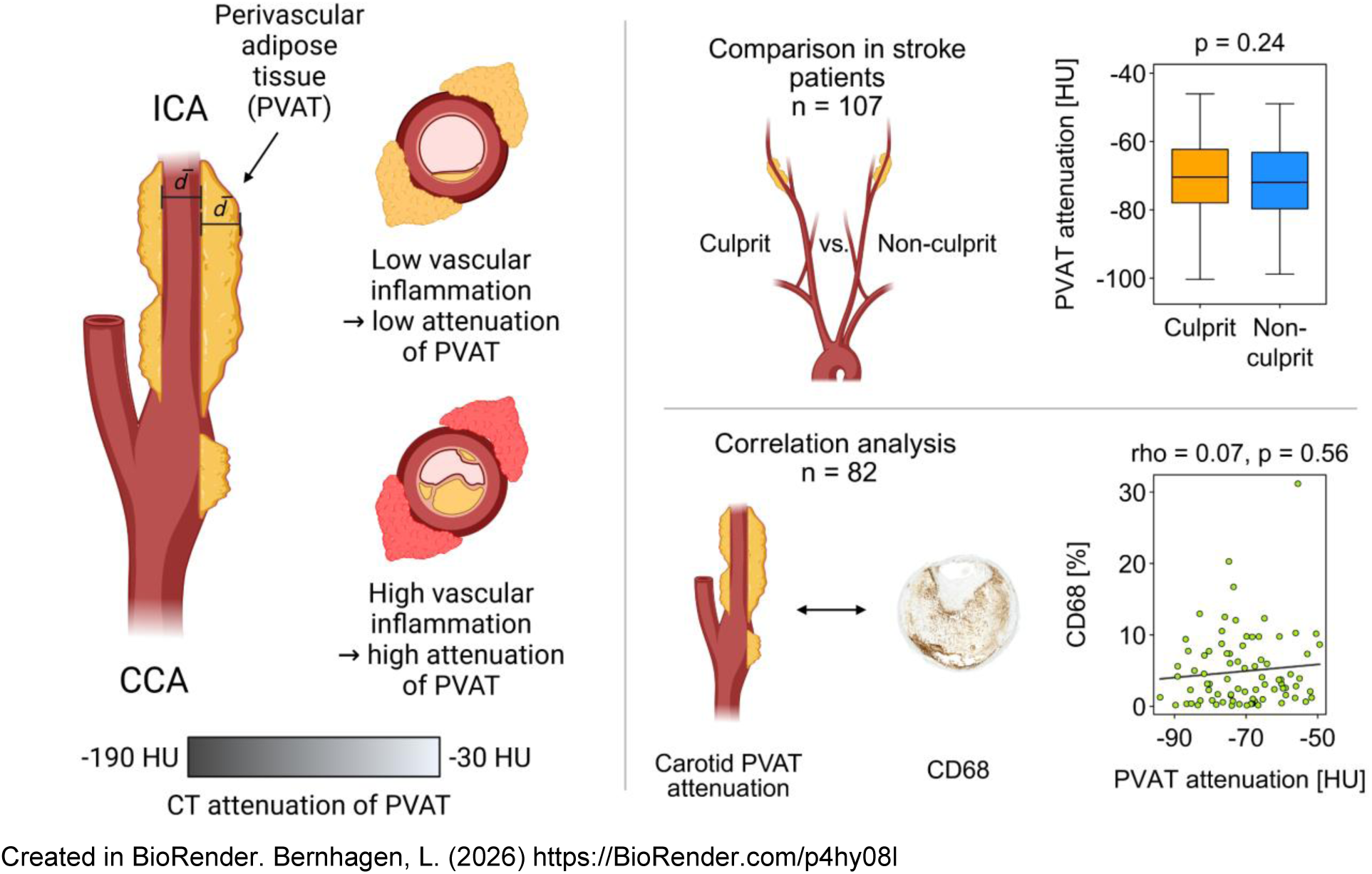

## Introduction

Atherosclerotic cardiovascular disease remains the main cause of mortality and morbidity worldwide.^1^ Inflammation is now recognized as a central driver of atherosclerosis,^2^ with large-scale trials demonstrating that anti-inflammatory therapies like colchicine^3,4^ or canakinumab^5^ can reduce cardiovascular risk. However, specific biomarkers for vascular inflammation are lacking, which hinders patient selection for emerging anti-inflammatory therapies. While hsCRP has been assessed for clinical use,^6^ circulating markers largely reflect systemic rather than localized arterial inflammation, limiting their specificity.^7^ As a result, imaging biomarkers have gained interest for directly quantifying plaque inflammation.^8^ Positron emission tomography (PET) with 18F-fluordeoxyglucose (FDG), or newer tracers, is a promising tool, as carotid FDG uptake correlates with plaque macrophage content and recurrent stroke risk.^9–13^ Yet, PET faces limitations for routine use, including high radiation exposure, cost, and limited availability across hospitals.

Recently, perivascular adipose tissue (PVAT) has emerged as a potential readout of vascular inflammation.^14^ Inflammation increases PVAT density,^14^ which can be clinically measured via CT imaging, as tissue density determines CT attenuation. The mean Hounsfield unit (HU) of PVAT, termed the Fat Attenuation Index (FAI), has emerged as a biomarker for plaque inflammation.^14^ FAI has been widely studied in coronary artery disease, where an FAI > -70 HU in coronary CTA was predictive of higher all-cause and cardiac mortality.^15^ Whether these findings generalize to other vascular beds remains unclear. In carotid atherosclerosis, early studies report higher FAI in symptomatic versus asymptomatic patients^16,17^ and associations with intraplaque hemorrhage.^18^ Despite these promising results, standardized approaches to FAI assessment are lacking, as many studies rely on a single CT slice^16,18^ with risk of high observer variability. Importantly, histologic validation of carotid FAI against plaque inflammation is currently lacking.

Here, we sought to address these gaps by developing a standardized method for carotid PVAT quantification on CTA and by evaluating its clinical and biological relevance. Specifically, we assessed whether carotid PVAT attenuation was associated with symptomatic culprit atherosclerotic lesions and whether it related to histological and molecular features of plaque inflammation and vulnerability in carotid endarterectomy specimens.

## Methods

### Study design overview

For the purposes of the current study, we analyzed carotid CTA scans from several cohorts. We first developed a standardized pipeline for PVAT segmentation and quantification of attenuation in the common carotid and internal carotid arteries. We then evaluated PVAT metrics in patients from the AtherOMICS, DEMDAS/DEDEMAS, and PROSCIS studies (n=147) with ischemic stroke or transient ischemic attack attributed to carotid stenosis, using both intrapatient comparisons (culprit versus contralateral carotid artery) and interpatient comparisons (culprit carotid artery versus control arteries without symptomatic extracranial carotid disease). Finally, leveraging the AtherOMICS cohort of patients undergoing carotid endarterectomy (n=117), we examined associations between preoperative PVAT metrics and histological and molecular features of plaque inflammation and vulnerability, including macrophage content, fibrous cap thickness, and plaque levels of inflammatory cytokines.

### Study cohorts

The AtherOMICS biobank^19^ is a single-center study including patients with atherosclerotic carotid artery stenosis undergoing carotid endarterectomy at LMU University Hospital, Munich, Germany. Both symptomatic patients with ischemic stroke, transient ischemic attack, retinal artery occlusion, or amaurosis fugax, and asymptomatic patients are enrolled. After the surgery, the carotid plaque sample is collected and stored for further analyses. Carotid plaque tissue and clinical data are collected prospectively. Recruitment started in 2022 and is ongoing. For the intrapatient and interpatient analysis, all 77 symptomatic patients recruited through May 2024 were included, and for the histological and molecular correlation analysis, all 117 symptomatic and asymptomatic patients recruited through the same date were included.

The DEDEMAS (2011-2013, single-center) and DEMDAS (2013-2018, multicenter across six tertiary stroke centers in Germany) studies are observational cohorts designed to investigate predictors of cognitive decline after acute stroke (ClinicalTrials.gov Identifier: NCT01334749). Details about the study inclusion and exclusion criteria have been previously described.^20^ Among 355 patients recruited at our center (Institute for Stroke and Dementia Research, LMU University Hospital, Munich, Germany), 35 patients with acute ischemic stroke attributed to extracranial carotid artery stenosis were included in the intrapatient and interpatient analyses. For interpatient comparison, a randomly selected sample of 13 patients with ischemic stroke not attributed to extracranial carotid atherosclerosis were analyzed as a control group.

PROSCIS is a two-center observational cohort conducted in Berlin and Munich, initiated in 2010 and 2011, respectively, aiming to develop risk prediction models after first-ever stroke (ClinicalTrials.gov Identifier: NCT01364168). The inclusion and exclusion criteria have been previously described.^21^ Among 466 patients recruited at our center through July 2023, 35 patients with stroke attributed to extracranial carotid artery stenosis were included in the intrapatient and interpatient analyses.

Patients were excluded from the current analyses if CTA data were unavailable, images were of substantially low quality, or if segmentation was precluded by prior carotid stenting or total artery occlusion.

Data for this investigation were sourced from primary studies conducted in accordance with the Declaration of Helsinki, with ethical approval from their respective local ethics committees and informed consent obtained from all participants. The current analysis protocol was approved by the Ethics Committee of the Medical Faculty of LMU Munich (Protocol No. 22-0135).

### Computed tomography angiography (CTA)

#### Data acquisition

For all symptomatic patients meeting the inclusion criteria for this study, we extracted clinical CTA scans performed during the initial routine acute stroke workup. For asymptomatic AtherOMICS patients, we extracted the most recent CTA scan obtained prior to endarterectomy, often acquired in outpatient settings outside our hospital. CT models and protocols varied across patients.

#### Image analysis

For segmentation in the carotid CTA scans, we used the open-source 3D Slicer software^22,23^ (version 5.6.2). To standardize the procedure and capture regions commonly affected by plaque, we first segmented 2 cm distal from the carotid bifurcation for the internal carotid artery (ICA) and 1 cm proximal for the common carotid artery (CCA). We generated an initial vessel segmentation along these 3 cm segments using a threshold of ≥ 100-300 HU (exact value customized per CTA scan to enable high contrast of lumen and calcified plaque to the surrounding tissue) and then manually expanded it to incorporate soft plaque and the outer arterial wall. We also analyzed the 2-cm-ICA segment separately to allow region-specific PVAT assessment.

Subsequently, we segmented PVAT, defined as all voxels between -190 and -30 HU,^14,24^ within a radius corresponding to the mean vessel diameter^14^ determined using the “Extract centerline” function of the VMTK extension of 3D Slicer.^25^ We then calculated the FAI-PVAT as the mean HU of all voxels in the PVAT segment. To account for variation in CTA acquisition protocols, we normalized FAI-PVAT values^26^ to a kilovoltage peak of 90 kV (except for paired analyses). We excluded voxels with attenuation values between -190 and -30 HU in the vicinity of the trachea to avoid misclassification as fat due to CT partial volume effects arising from adjacent air and soft tissue within the same voxel.

Beyond FAI-PVAT, we further computed relative FAI metrics to account for additional factors affecting PVAT attenuation, including BMI^14,27^ and CT scanner settings.^28,29^ Relative metrics help compare between patients in interpatient analyses, but also control for local confounding factors in intrapatient analyses. These included: the volumetric perivascular characterization index (VPCI), defined as the percentage decrease from FAI-PVAT to the outermost 1 mm thick adipose tissue ring at a distance of 19 to 20 mm from the carotid artery;^14^ the FAI-slope, defined as the FAI gradient from inner to outer 1 mm adipose tissue rings across a distance of 20 mm from the carotid artery and quantified as the linear regression coefficient of distance on FAI;^14^ and the FAI-hCCA, defined as the percentage decrease from FAI-PVAT in the ICA and CCA segments to a CCA segment 2 cm proximal to the bifurcation, which we used as a presumed healthy reference (hCCA = healthy CCA). This reference segment measured 2 cm in length.

### Plaque analyses in AtherOMICS

#### Plaque biobanking

In the context of the AtherOMICS biobank,^19^ carotid plaque specimens are collected directly from the operating room after endarterectomy within a median of 40 minutes [IQR 33 to 50 minutes] after excision. As part of a standardized protocol,^19^ each plaque is divided into a main segment containing the site of maximal stenosis for histological analyses and adjacent segments for further molecular and cellular profiling. The main segment is fixated in formaldehyde for 24 hours, stored in 70% ethanol after washing under water for 1 hour, and is finally decalcified in aTris-buffered 200 mM EDTA solution at a pH of 8.0 at 4°C.^30^ The duration of decalcification is adjusted to the amount of calcification. The remaining segments are flash-frozen in 2-methylbutane cooled with liquid nitrogen and stored at -80°C for molecular analyses.

#### Histology and image analysis

After the initial processing, the main decalcified plaque sample is embedded in paraffin and sectioned into 1 mm slices. Serial sections of 5µm thickness are taken that undergo stainings with H&E, anti-alpha-smooth-muscle-actin (ɑSMA) immunohistochemistry (IHC) for smooth muscle cells and anti-CD68 IHC for macrophages using 3,3’-diaminobenzidine (DAB), and Picrosirius Red for collagen. Slides are scanned at high resolution with a microscope (Carl Zeiss NTS Ltd., Oberkochen, Germany) and the resulting whole-slide images are stored for digital processing. To analyze the images, we used QuPath (version 0.4.4).^31^

After importing every image of each sample, we quantified macrophage content by applying the color deconvolution method^32^ to extract DAB staining. This was accomplished by extracting the staining vectors from a small representative rectangular area in one exemplary image. Then, we created a pixel thresholder to include all pixels with a DAB optical density above a certain value. Additionally, to measure the total pixel count of every tissue section independent of staining (by removing background pixels) we applied another threshold, based on the average RGB value. To calculate the relative CD68 amount at the plaque level we divided the sum of DAB pixels of all images by the total tissue pixel count of all images for each sample.

We measured fibrous cap thickness in Sirius Red staining, using only sections that contained at least half of the plaque circumference. Fibrous cap thickness was defined as the minimal rectangular distance between luminal surface and the underlying lipid core.

#### Plaque cytokine levels

Using plaque segments adjacent to the main segment, we performed cell lysis with a RIPA lysis buffer and complete Mini protease inhibitor cocktail (Sigma-Aldrich, St. Louis (MO), USA). We visually tested protein degradation with SDS-PAGE and subsequent Coomassie Blue staining. Proteomic profiling was performed on 96-well PCR plates using Olink’s Explore 3072 panel, a proximity extension assay platform that utilizes DNA-conjugated antibodies and subsequent DNA amplification and sequencing to determine protein abundance.^33^ The protein concentration is represented as normalized protein expression (NPX), calculated from the resulting Ct values and adjusted for inter-plate variation. For the purposes of the current analyses, we extracted plaque IL-6 and TNF levels that have been previously correlated with PVAT metrics in coronary atherosclerosis.^14,34^

### Statistical analysis

We performed all statistical analyses using R (version 4.4.1)^35^ including the stringr,^36^ pwr^37^ and DescTools^38^ packages and RStudio (version 2024.4.2.764).^39^ For intrapatient analyses, we compared PVAT attenuation metrics between culprit and contralateral carotid arteries using paired, two-sided t-tests. For interpatient analyses, we compared PVAT metrics from culprit carotid arteries of patients with carotid atherosclerosis to the randomly chosen right or left carotid arteries of stroke patients without extracranial carotid atherosclerosis using unpaired, two-sided t-tests. We assessed associations between

PVAT metrics and histological and molecular markers of plaque inflammation and vulnerability (CD68, fibrous cap thickness, IL-6, and TNF) using Spearman’s rank correlation coefficients. All tests were two-sided, and we considered p-values < 0.05 statistically significant.

## Results

### Description of patient cohorts

We performed two main levels of analyses: (1) comparisons of PVAT metrics between culprit carotid lesions associated with clinical events and non-culprit carotid segments, either within patients (contralateral arteries) or, secondarily, in external controls; and (2) correlations of PVAT metrics with histological and molecular features of plaque inflammation and vulnerability. A flowchart of patient selection is shown in **Figure 1**. For the comparative analyses, we included 147 patients with symptomatic carotid artery disease in the DEDEMAS/DEMDAS (n=35), PROSCIS (n=35), and AtherOMICS cohorts (n=77). After excluding patients due to unavailable CTA data, segmentation constraints (total occlusion or stenting), or general low image quality, 107 patients remained for intrapatient comparisons between culprit and contralateral carotid arteries. In secondary analyses, we compared these patients (and four more patients, where only the culprit side could be analyzed because of stenting or total occlusion in the non-culprit carotid artery) to 13 controls from the DEMDAS cohort with ischemic stroke but no evidence of extracranial carotid atherosclerosis. For the correlations with plaque inflammation and vulnerability, we included 117 patients from AtherOMICS undergoing endarterectomy due to symptomatic or asymptomatic carotid disease. After exclusions, 90 patients remained for analyses.

**Figure 1:**
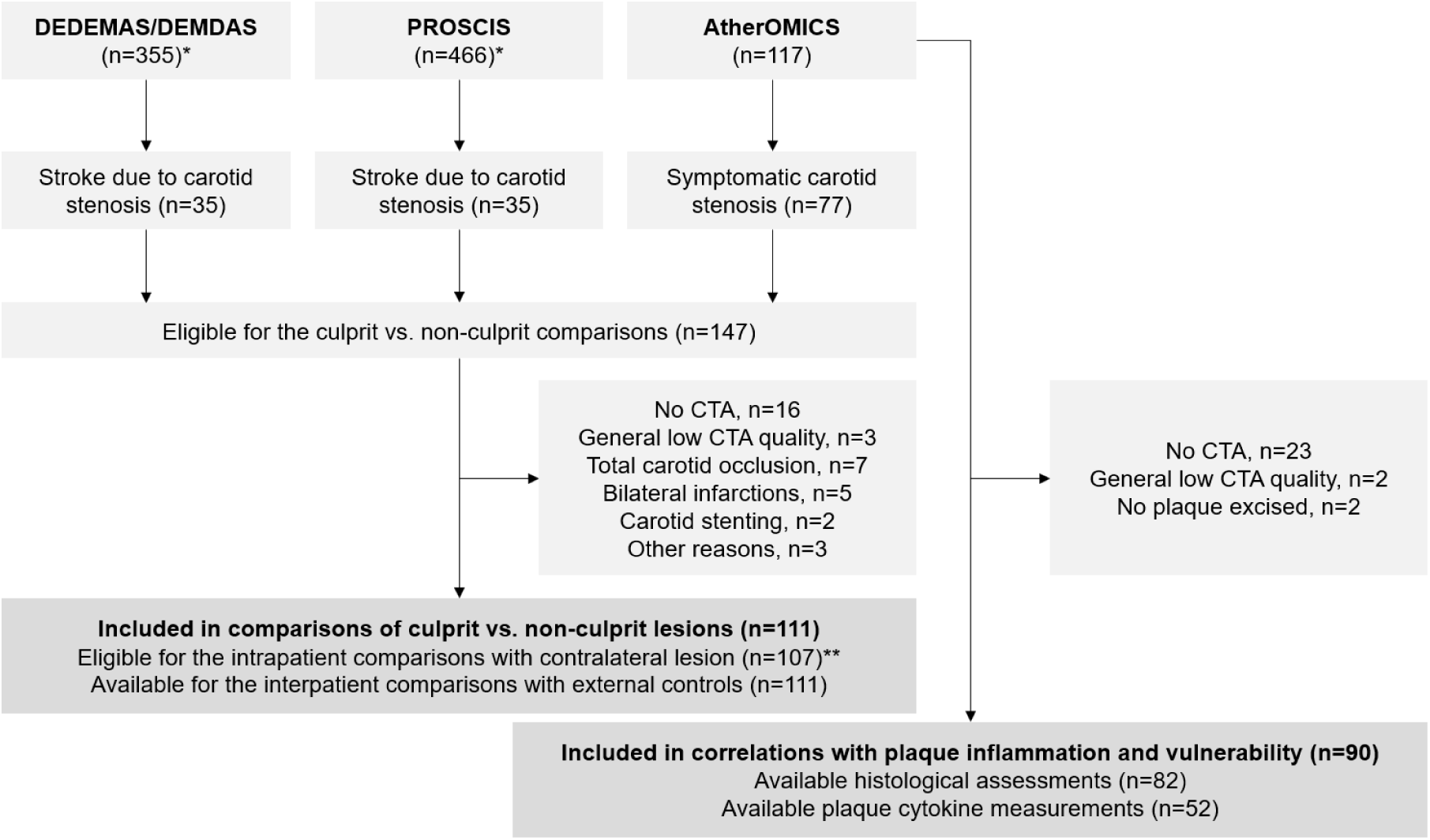
Flowchart of the study cohorts *Inclusion was restricted to patients recruited at the LMU Klinikum study site. **Four additional patients were excluded from the paired comparison of culprit versus the contralateral carotid arteries (intrapatient analysis) due to segmentation hurdles on the non-culprit side; two patients were excluded due to carotid stenting and two due to a non-segmentable occlusion.

The baseline characteristics of patients included in the two analyses are presented in **Table 1**. In the comparative analyses, mean age of the 111 included patients was 75.3 years (SD 8.8) and 69 of them (62.2%) were male. As expected, this was a cohort with high burden of cardiovascular risk factors with 67.3%, 24.8%, and 51.5% having a history of hypertension, diabetes mellitus, and dyslipidemia, respectively. The mean BMI was 26.2 kg/m^2^ (SD 4.3) and 62.7% of study participants were current or former smokers. Among 90 patients included in the correlation analyses with plaque inflammation and vulnerability, the mean age was 73.7 years (SD 9.1) and 60 were male (66.7%). Similarly, this cohort exhibited a high burden of cardiovascular risk factors, with comparable prevalence of hypertension, diabetes mellitus, dyslipidemia, and smoking. Patients excluded from the comparative analyses were younger than the included patients, whereas patients excluded from the correlation analyses had significantly higher rates of history of high LDL cholesterol; otherwise, there were no significant differences between eligible participants included in the analyses and participants excluded for both cohorts (**Figure 1** and **Supplementary Table 1**). The baseline characteristics of the randomly selected stroke controls without carotid atherosclerotic disease for the interpatient analysis are presented in **Supplementary Table 2**. Regarding CTA parameters (**Supplementary Table 3**), slice thickness varied across scans, but it was 1 mm or less for >79% of the analyzed cases. The kilovoltage peak (kVp) was in the range between 70 and 120 kV and most CTAs (>81%) were performed on scanners manufactured by Siemens Healthineers (Erlangen, Germany).

**Table 1:** Baseline characteristics

| Variable | Culprit vs. Non-culprit analysis | n | Correlation analysis | n |
| --- | --- | --- | --- | --- |
| n | 111 |  | 90 |  |
| Age (years, mean ± SD) | 75.3 (± 8.8) | 111 | 73.7 (± 9.1) | 90 |
| Sex (male), N (%) | 69 (62.2) | 111 | 60 (66.7) | 90 |
| BMI (kg/m<sup>2</sup>, mean ± SD) | 26.2 (± 4.3) | 110 | 26.4 (± 4.5) | 89 |
| History of hypertension, N (%) | 74 (67.3) | 110 | 67 (75.3) | 89 |
| History of diabetes mellitus, N (%) | 27 (24.8) | 109 | 24 (27.3) | 88 |
| Former smoker, N (%) | 38 (34.5) | 110 | 32 (36.8) | 87 |
| Current smoker, N (%) | 31 (28.2) | 110 | 25 (28.7) | 87 |
| History of high LDL cholesterol, N (%) | 52 (51.5) | 101 | 57 (69.5) | 82 |
| History of cardiovascular disease, N (%) | 43 (43.9) | 98 | 51 (67.1) | 76 |
| Antiplatelet, N (%) | 64 (59.3) | 108 | 62 (69.7) | 89 |
| Anticoagulation, N (%) | 16 (14.8) | 108 | 20 (22.5) | 89 |
| Statins and other lipid lowering therapy, N (%) | 56 (51.9) | 108 | 66 (74.2) | 89 |
| Antihypertensive medication, N (%) | 73 (67.6) | 108 | 71 (79.8) | 89 |
| Antidiabetics, N (%) | 24 (22.2) | 108 | 21 (23.6) | 89 |

SD = standard deviation. BMI = body mass index.

### Quantification of PVAT metrics

We computed FAI-PVAT and three relative PVAT attenuation metrics (VPCI, FAI-slope, and FAI-hCCA) for the distal CCA, proximal ICA, and their composite across a total of 148 patients (**Figure 2**). PVAT volume defined as voxels between -190 and -30 HU surrounding a radius equal to the artery’s diameters varied across individuals. The distributions of the 4 metrics are shown in **Supplementary Figure 1**. As expected, we observed moderate to strong correlations between FAI-PVAT and relative PVAT metrics (**Supplementary Figure 2**). FAI values showed expected gradients with increasing distance from the vessel wall, consistent with prior observations in coronary arteries^14^ (**Supplementary Figure 3**). Some level of image artifacts related to dental protheses (or in a few cases due to motion during the CTA procedure) were present in 68 of the analyzed cases (46%) and influenced all FAI parameters, decreasing FAI-PVAT, VPCI and FAI-hCCA and increasing FAI-slope (**Supplementary Table 4**). When examining relationships with age, sex, and BMI, we found no significant associations across the PVAT metrics (**Supplementary Figure 4**). Among technical parameters, we found no evidence that slice thickness, kVp, and scanner manufacturer significantly influence the PVAT metrics (**Supplementary Figure 5**).

**Figure 2:**
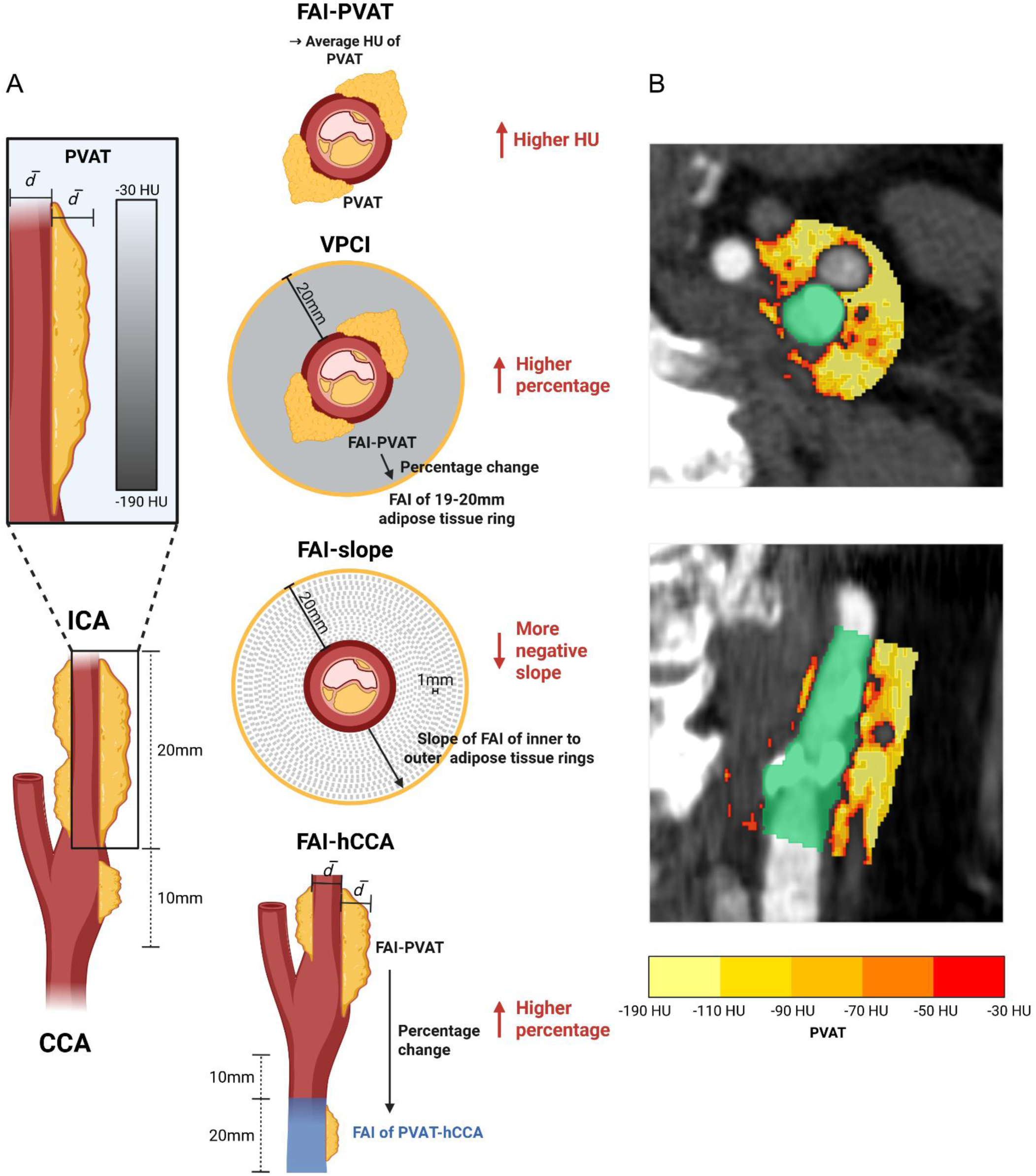
Overview of the fat attenuation index (FAI) parameters (A) Perivascular adipose tissue (PVAT) is defined as all voxels with Hounsfield Units (HU) between -30 and -190, within a distance around the carotid artery corresponding to the mean vessel segment diameter. From the internal carotid artery (ICA) the proximal 20 mm are analyzed and from the common carotid artery (CCA) the distal 10 mm. For the FAI-PVAT analysis the average HU of the PVAT was calculated. The volumetric perivascular characterization index (VPCI) represents the percentage decrease from the FAI-PVAT to the FAI of a 1 mm thick adipose tissue ring, in a distance of 19-20 mm from the outer vessel wall. The FAI-slope was calculated as the slope of the linear regression of the FAI from the inner to the outer 1 mm thick ring from 0 to 20 mm from the outer vessel edge. The FAI-hCCA (healthy CCA) shows the percentage decrease from the FAI-PVAT to the FAI of a CCA segment, 20 mm proximal to the bifurcation, presumed to be a healthy reference. The red arrows indicate the hypothesized change in the FAI parameters with more carotid inflammation. (B) Axial and coronal CTA view of the FAI-PVAT analysis. The ICA and CCA segment is presented in green, the PVAT in red (higher HU), orange and yellow (lower HU). Created in BioRender. Bernhagen, L. (2026) https://BioRender.com/ydsxugd

### Comparisons of PVAT metrics between culprit and non-culprit carotid plaques

We then compared PVAT metrics in the culprit lesion versus the contralateral carotid segment among patients with symptomatic carotid disease in paired intrapatient analyses. Across 107 patients, we found no significant differences across the PVAT metrics either when analyzing ICA and CCA as a composite segment or ICA separately (**Figure 3 and Supplementary Figure 6**). FAI-PVAT, and all relative metrics (VPCI, FAI-slope, FAI-hCCA) were largely similar in culprit lesions associated with symptomatic clinical events and contralateral non-culprit lesions. These results were similar in sensitivity analyses, where FAI-PVAT and FAI-hCCA were computed based on a standard 5 mm radius instead of adjusted to the radius of the index vessel (**Supplementary Figure 7**), as well as when pursuing analyses excluding any individuals with artifacts (**Supplementary Figure 8**).

**Figure 3:**
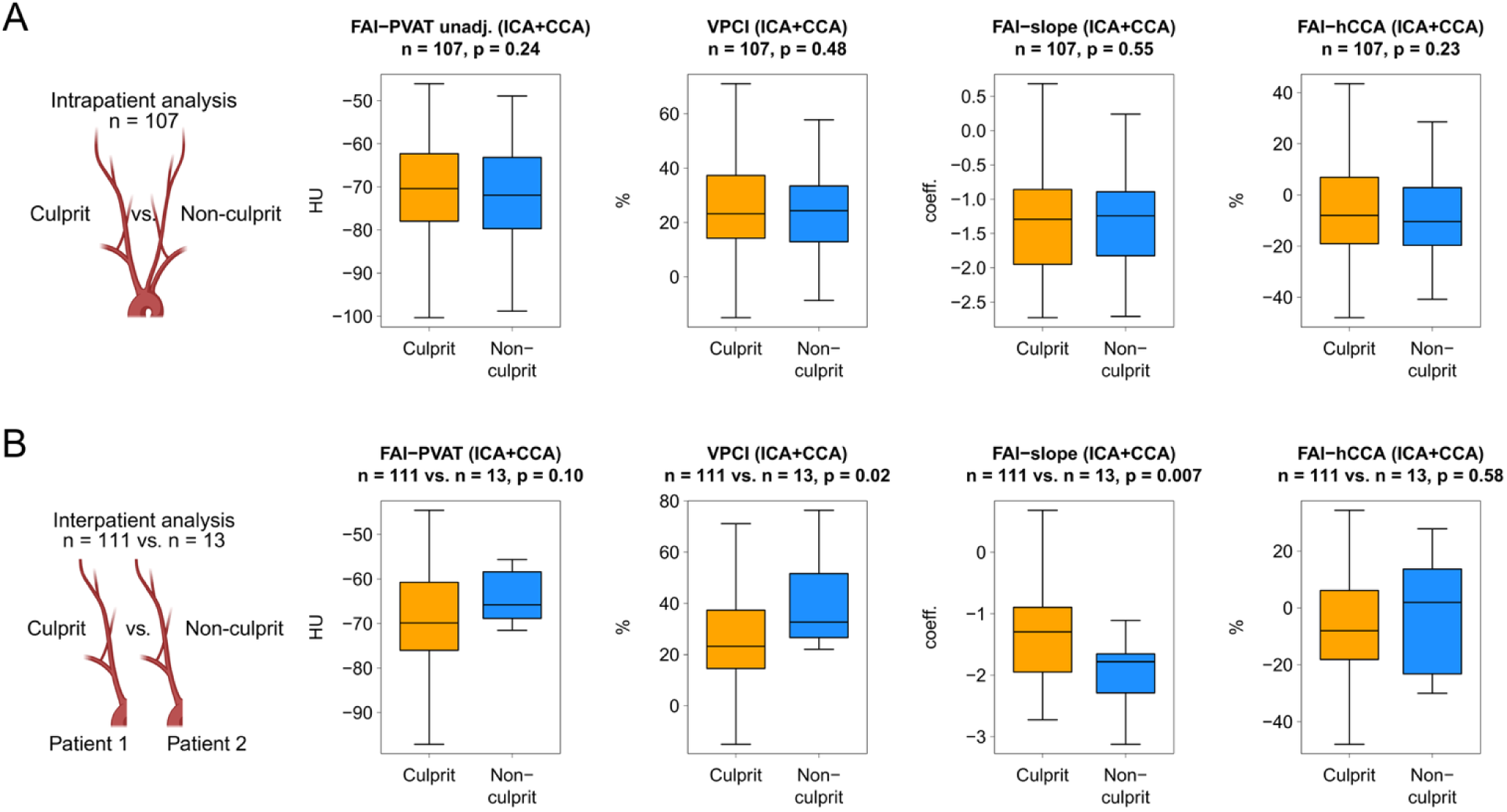
Comparison of fat attenuation index parameters in culprit versus non-culprit carotid arteries (A) Paired comparison of the fat attenuation index (FAI) parameters between the culprit and the contralateral non-culprit side of 107 patients with stroke (Intrapatient analysis). (B) Unpaired comparison of the FAI of the culprit carotid artery of 111 symptomatic patients and a random non-culprit side of 13 stroke patients with non-carotid stroke etiology (Interpatient analysis). In the latter analysis the FAI-PVAT was adjusted by the kilovoltage peak (kVp) of each patient’s CTA scan. The analyzed vessel segments include 2 cm of the internal carotid artery (ICA) and 1 cm of the common carotid artery (CCA) around the bifurcation. PVAT = perivascular adipose tissue. VPCI = volumetric perivascular characterization index. hCCA = healthy common carotid artery. HU = Hounsfield units. unadj. = unadjusted. Created in part with BioRender. Bernhagen, L. (2026) https://BioRender.com/gchun17 and https://BioRender.com/yi80xa4

Similarly, we found no evidence for higher PVAT attenuation among culprit lesions, in the interpatient analyses comparing 111 culprit lesions to 13 vessels without evidence of atherosclerosis from control patients. As opposed to the original hypothesis, VPCI was higher and FAI-slope lower in segments from ICA and CCA without evidence of atherosclerosis (**Figure 3**). Similar trends were observed when focusing only on the ICA segment (**Supplementary Figure 6**), when computing FAI-PVAT, VPCI and FAI-hCCA based on a fixed 5 mm radius (**Supplementary Figure 7**) as well as in sensitivity analyses excluding patients with image artifacts (**Supplementary Figure 8**).

### Correlations between PVAT metrics and plaque inflammation and vulnerability

Subsequently, using paired data CTA imaging and histological and molecular analyses of plaque tissue, we explored correlations of PVAT metrics with markers of plaque inflammation and vulnerability in patients from the AtherOMICS cohort. Our analyses revealed no significant correlations between macrophage content, as captured by CD68 staining and any of the four PVAT metrics (n=82, **Figure 4**) quantified for the combined ICA and CCA segment (r ranging from -0.11 to 0.07, all p >0.03). Our analyses revealed inverse correlations to what we would expect between PVAT metrics and fibrous cap thickness, with higher PVAT attenuation being associated with a thicker fibrous cap, which is a marker of a stabler plaque (n=49, **Figure 4**). Finally, we found no significant correlation between PVAT metrics and plaque protein levels of IL-6 or TNF, as quantified with the Olink Explore platform (n=52, **Figure 5**). These results were largely stable in sensitivity analyses for FAI-PVAT, as well as when excluding participants without artifacts from the analyses (**Supplementary Figure 8**).

**Figure 4:**
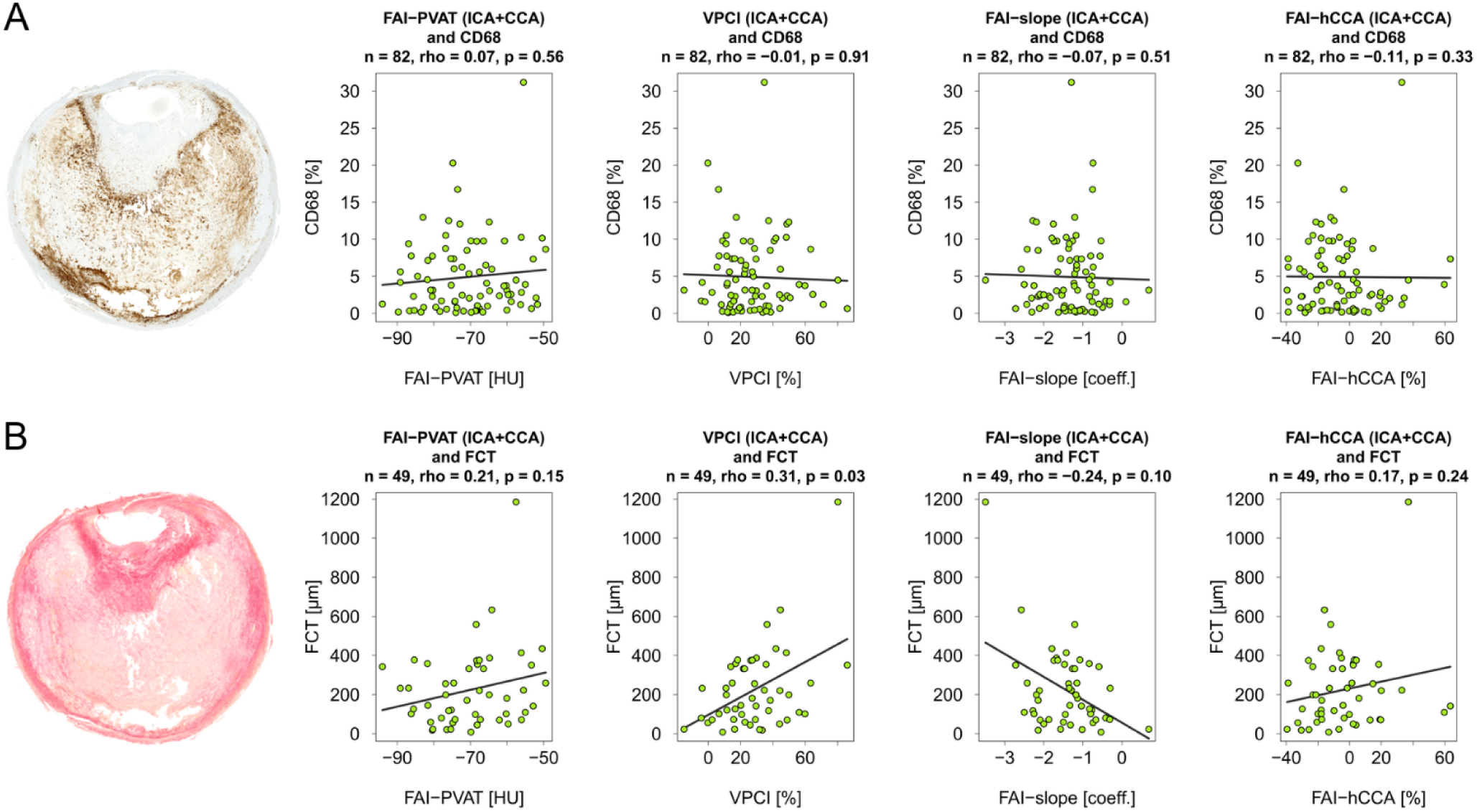
Correlation analysis of fat attenuation index parameters and histological features Spearman’s rank correlation was used to assess the relationship between fat attenuation index (FAI) parameters and CD68 amount (A) and fibrous cap thickness (B) in culprit carotid arteries of stroke patients. CD68 is predominantly expressed by macrophages and commonly used to quantify plaque inflammation. Fibrous cap thickness describes the width of the connective tissue separating the lipid core from the arterial lumen, whereas a thicker cap is associated with more plaque stability. The FAI results are shown for a bifurcation segment comprising of 2 cm of the internal carotid artery (ICA) and 1 cm of the common carotid artery (CCA). For histological analysis the main part of the ICA specimens containing the maximum stenosis was used. PVAT = perivascular adipose tissue. VPCI = volumetric perivascular characterization index. hCCA = healthy common carotid artery. FCT = fibrous cap thickness. HU = Hounsfield units.

**Figure 5:**
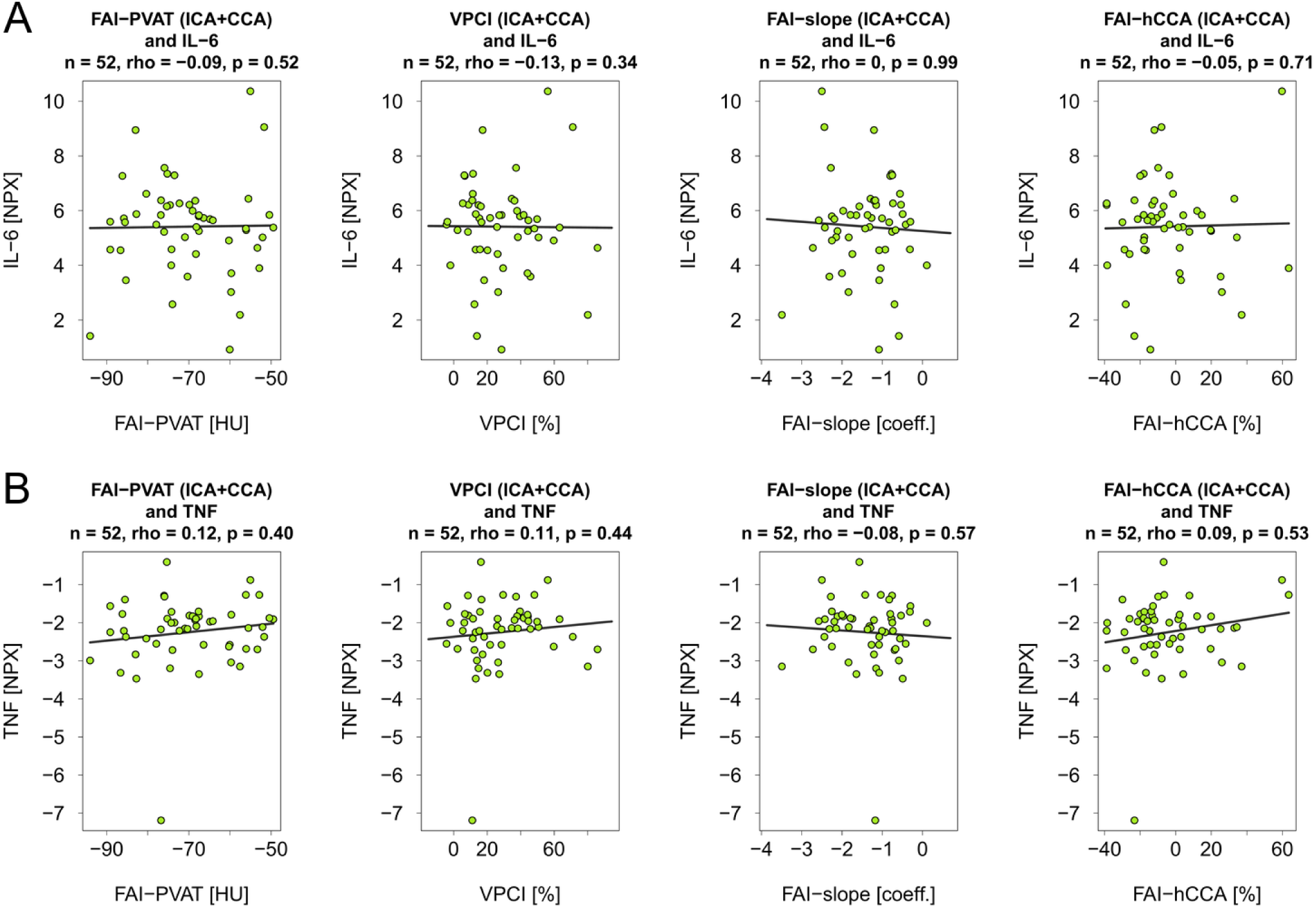
Correlation analysis of fat attenuation index parameters and proteomic markers The relationship of fat attenuation index (FAI) parameters and IL-6 (A) and TNF (B) is presented utilizing Spearman’s rank correlation. For the FAI results, the analyzed vessel segment consists of 2 cm of the internal carotid artery (ICA) and 1 cm of the common carotid artery (CCA) around the bifurcation. Proteomic analyses were performed on a peripheral part of the carotid specimens. PVAT = perivascular adipose tissue. VPCI = volumetric perivascular characterization index. hCCA = healthy common carotid artery. IL-6 = interleukin 6. TNF = Tumor necrosis factor. HU = Hounsfield units. NPX = normalized protein expression.

To ensure that we had sufficient statistical power to detect a significant correlation between PVAT metrics and CD68 content, we estimated that we could detect any r > 0.3 or < -0.3 as statistically significant with our sample size of n=82 (1-β=0.8, α=0.05). This is well within the range previously reported for correlations of CD68 content with FDG uptake in PET imaging that has been proposed as a marker of local inflammation (range of r in previous studies between 0.55^40^ and 0.85^41^).

## Discussion

In this study, we developed and applied a standardized pipeline for quantifying PVAT attenuation around the carotid artery on routine CTA. Contrary to findings previously reported in the coronary circulation, we observed no systematic differences in PVAT attenuation metrics between culprit and non-culprit carotid plaques, whether comparing culprit to contralateral non-culprit lesions within patients with symptomatic carotid atherosclerosis, or comparing culprit lesions to carotid arteries of patients without evidence of symptomatic atherosclerosis. Moreover, CTA-derived PVAT attenuation metrics showed no convincing correlation with histological markers of plaque inflammation and stability (macrophage content, fibrous cap thickness) or with plaque protein levels of the inflammatory cytokines IL-6 and TNF in surgically excised carotid specimens.

Our study extends the literature examining whether the concept of PVAT attenuation as a marker of vascular inflammation, originally established in the coronary circulation, can be transferred to the carotid arteries. While our findings may appear to contradict earlier reports of significant differences in FAI between culprit and non-culprit carotid arteries,^16,17^ these studies quantified FAI using a single, manually placed region of interest in the pericarotid fat, an approach that is inherently more susceptible to observer variability and lacks the standardization of volumetric, vessel-adapted segmentation. In contrast, another study employing a methodological approach similar to ours also found no significant difference between culprit and non-culprit carotid FAI,^42^ consistent with our results. Separately, early studies have reported associations between carotid FAI and recurrent cerebrovascular events,^43,44^ raising the possibility that pericarotid fat density may hold prognostic value even in the absence of a detectable culprit-lesion signal. However, these findings warrant validation in larger, prospective cohorts.

Most importantly, our study provides no evidence supporting PVAT attenuation reflect carotid plaque inflammation or vulnerability at the histological or molecular level. In the coronary arteries, a close paracrine relationship between atherosclerotic plaque and PVAT has been proposed as the biological basis for using PVAT attenuation as a surrogate marker of plaque inflammation.^14^ A preclinical study^14^ reported that the differentiation of human preadipocytes was inhibited after exposure to inflammatory cytokines released by the atherosclerotic plaque, like IL-6 and TNF, leading to a reduction of lipid content, which can be detected by CT imaging. In contrast, our paired analysis of CTA and endarterectomy specimens showed no correlation between any CTA-derived PVAT attenuation metric and macrophage content, fibrous cap thickness, or plaque levels of IL-6 and TNF. This lack of association, observed in a cohort with adequate statistical power to detect moderate correlations, raises questions about whether PVAT attenuation metrics can be generalized as markers of plaque inflammation beyond the coronary circulation.

Our results are subject to several limitations. First, CTA images from many patients were affected by streak artifacts caused by dental implants. To our knowledge, no study has systematically examined the effect of such artifacts on FAI measurements, but because they can generate Hounsfield units within the range attributed to adipose tissue (-190 to - 30 HU), the resulting disruption of PVAT quantification may be substantial. However, we consider it unlikely that streak artifacts may have substantially influenced our main conclusions, as the results were largely consistent in sensitivity analyses excluding patients with artifacts. Second, we used routine CTA scans, which were acquired on different CT devices with non-standardized parameters. Despite kVp adjustment, this heterogeneity likely renders FAI-PVAT measurements imprecise in unpaired analyses, as additional scanner-related factors beyond kVp are known to influence Hounsfield units.^29^ Third, for symptomatic patients we predominantly used CT scans acquired shortly after an acute ischemic event, and whether and how this timing might affect FAI metrics remains unclear. Fourth, while we aimed to standardize our approach, the step of carotid vessel segmentation relied on both manual and semi-automated tools, which may limit the reproducibility of our image analysis pipeline. Fifth, we observed that perivascular fat around the carotid arteries appeared limited in volume for some patients, in contrast to the abundant epicardial adipose tissue surrounding the coronary arteries. This could disproportionately affect radius-based analyses, which include all fat within a fixed distance of the vessel. Finally, our histological analyses are subject to additional constraints. The extent of atherosclerotic plaque preserved in each specimen varied across patients, as undamaged plaque removal is not always surgically feasible and depends on the surgical technique used (thromboendarterectomy versus eversion endarterectomy), which may have affected the validity of CD68 and fibrous cap thickness measurements. Moreover, proteomic analyses were performed on a peripheral plaque segment rather than the main specimen, which was prioritized for histology, which may have influenced the representativeness of quantified IL-6 and TNF levels.

## Conclusion

In contrast to the coronary arteries, carotid PVAT attenuation on CTA was not associated with symptomatic culprit lesions, nor with histological or molecular markers of plaque inflammation and vulnerability in patients with symptomatic carotid atherosclerosis. These findings question the transferability of this CT-based marker of arterial inflammation to extracoronary atherosclerosis and warrant further investigation.

## Acknowledgements

We are grateful to all study participants for their contributions to AtherOMICS, DEMDAS/DEDEMAS, and PROSCIS. We also thank Jana Mattar, Paulo Vinicius Gil Alabarse, Mohamad Ali Antabi, Panagiotis Zangas, Iulia Lupul and Anastasiia Osadcha who contributed to the study operations of AtherOMICS.

## Data availability statement

Due to ethical and data protection constrains, individual-level data cannot become publicly available. Requests for data access together with a research proposal can be directed to the corresponding author and will be reviewed. Access is subject to scientific review and completion of a material transfer agreement with the LMU Klinikum.

## Author Information

DEMDAS study group:

Martin Dichgans (Institute for Stroke and Dementia Research (ISD), University Hospital, LMU Munich, 81377 Munich, Germany; German Center for Neurodegenerative Diseases (DZNE), Munich 81377, Germany),

Matthias Endres (Department of Neurology with Experimental Neurology, Charité - Universitätsmedizin Berlin, Berlin, Germany; German Center for Neurodegenerative Diseases (DZNE), Berlin 10117, Germany; Center for Stroke Research Berlin (CSB), Charité - Universitätsmedizin Berlin, Berlin, Germany; German Centre for Cardiovascular Research (DZHK), partner site Berlin, Berlin, Germany; German Center for Mental Health (DZPG), partner site Berlin, Berlin, Germany),

Gabor C. Petzold (German Center for Neurodegenerative Diseases (DZNE), Bonn 53127, Germany; Department of Vascular Neurology, University Hospital Bonn, Bonn 53127, Germany),

Inga Zerr (Department of Neurology, University Medical Center Göttingen, Göttingen 37075, Germany; German Center for Neurodegenerative Diseases (DZNE), Göttingen 37075, Germany),

Michael Görtler (Department of Neurology, University Hospital, Otto-von-Guericke University Magdeburg, Magdeburg 39120, Germany; German Center for Neurodegenerative Diseases (DZNE), Magdeburg 39120, Germany),

Silke Wunderlich (Department of Neurology, School of Medicine and Health, Technical University of Munich, Munich, Germany),

Annika Spottke (German Center for Neurodegenerative Diseases (DZNE), Bonn 53127, Germany)

