## Supplemental Material for "Perivascular adipose tissue attenuation in CT angiography and plaque inflammation in patients with carotid atherosclerosis"

#### Supplementary Tables

##### Supplementary Table 1: Comparison of baseline characteristics between included and excluded patients

| Culprit vs. Non-culprit analysis | Included | n | Excluded | n | p-value |
|---|---|---|---|---|---|
| n | 111 | | 36 | | |
| Age (years, mean ± SD) | 75.3 (± 8.8) | 111 | 69.4 (± 9.3) | 36 | **0.002** |
| Sex (male), N (%) | 69 (62.2) | 111 | 28 (77.8) | 36 | 0.09 |
| BMI (kg/m², mean ± SD) | 26.2 (± 4.3) | 110 | 26.7 (± 4) | 36 | 0.58 |
| History of hypertension, N (%) | 74 (67.3) | 110 | 27 (75.0) | 36 | 0.38 |
| History of diabetes mellitus, N (%) | 27 (24.8) | 109 | 7 (19.4) | 36 | 0.51 |
| Former smoker, N (%) | 38 (34.5) | 110 | 16 (44.4) | 36 | 0.29 |
| Current smoker, N (%) | 31 (28.2) | 110 | 11 (30.6) | 36 | 0.78 |
| History of high LDL cholesterol, N (%) | 52 (51.5) | 101 | 17 (48.6) | 35 | 0.77 |
| History of cardiovascular disease, N (%) | 43 (43.9) | 98 | 11 (31.4) | 35 | 0.20 |
| Antiplatelet, N (%) | 64 (59.3) | 108 | 17 (47.2) | 36 | 0.21 |
| Anticoagulation, N (%) | 16 (14.8) | 108 | 4 (11.1) | 36 | 0.58 |
| Statins and other lipid lowering therapy, N (%) | 56 (51.9) | 108 | 14 (38.9) | 36 | 0.18 |
| Antihypertensive medication, N (%) | 73 (67.6) | 108 | 24 (66.7) | 36 | 0.92 |
| Antidiabetics, N (%) | 24 (22.2) | 108 | 3 (8.3) | 36 | 0.06 |

| Correlation analysis | Included | n | Excluded | n | p-value |
|---|---|---|---|---|---|
| n | 90 | | 27 | | |
| Age (years, mean ± SD) | 73.7 (± 9.1) | 90 | 74 (± 7.7) | 27 | 0.84 |
| Sex (male), N (%) | 60 (66.7) | 90 | 21 (77.8) | 27 | 0.27 |
| BMI (kg/m², mean ± SD) | 26.4 (± 4.5) | 89 | 25.9 (± 4.3) | 26 | 0.61 |
| History of hypertension, N (%) | 67 (75.3) | 89 | 20 (76.9) | 26 | 0.86 |
| History of diabetes mellitus, N (%) | 24 (27.3) | 88 | 8 (30.8) | 26 | 0.73 |
| Former smoker, N (%) | 32 (36.8) | 87 | 10 (43.5) | 23 | 0.56 |
| Current smoker, N (%) | 25 (28.7) | 87 | 4 (17.4) | 23 | 0.27 |
| History of high LDL cholesterol, N (%) | 57 (69.5) | 82 | 20 (90.9) | 22 | **0.04** |
| History of cardiovascular disease, N (%) | 51 (67.1) | 76 | 11 (45.8) | 24 | 0.06 |
| Antiplatelet, N (%) | 62 (69.7) | 89 | 18 (72.0) | 25 | 0.82 |
| Anticoagulation, N (%) | 20 (22.5) | 89 | 9 (36.0) | 25 | 0.17 |
| Statins and other lipid lowering therapy, N (%) | 66 (74.2) | 89 | 21 (84.0) | 25 | 0.31 |
| Antihypertensive medication, N (%) | 71 (79.8) | 89 | 18 (72.0) | 25 | 0.41 |
| Antidiabetics, N (%) | 21 (23.6) | 89 | 7 (28.0) | 25 | 0.65 |

Two-sided t-tests were used for continuous variables and chi-squared tests for binary variables. SD = standard deviation. BMI = body mass index.

**Supplementary Table 2: Baseline characteristics of randomly selected control cohort for the unpaired comparison of culprit and non-culprit carotid arteries**

| **Variable** | **Control cohort** | **n** |
|---|---|---|
| n | 13 | |
| Age (years, mean ± SD) | 70.5 (± 6.5) | 13 |
| Sex (male), N (%) | 8 (61.5) | 13 |
| BMI (kg/m², mean ± SD) | 25.9 (± 4.1) | 13 |
| History of hypertension, N (%) | 10 (76.9) | 13 |
| History of diabetes mellitus, N (%) | 2 (15.4) | 13 |
| Former smoker, N (%) | 5 (38.5) | 13 |
| Current smoker, N (%) | 3 (23.1) | 13 |
| History of high LDL cholesterol, N (%) | 1 (7.7) | 13 |
| History of cardiovascular disease, N (%) | 1 (7.7) | 13 |
| Antiplatelet, N (%) | 5 (38.5) | 13 |
| Anticoagulation, N (%) | 0 (0.0) | 13 |
| Statins and other lipid lowering therapy, N (%) | 3 (23.1) | 13 |
| Antihypertensive medication, N (%) | 5 (38.5) | 13 |
| Antidiabetics, N (%) | 2 (15.4) | 13 |

SD = standard deviation. BMI = body mass index.

**Supplementary Table 3: CTA parameters**

| | Culprit vs. non-culprit analysis | Unpaired culprit vs. non-culprit analysis (controls) | Correlation analysis |
|---|---|---|---|
| **Slice Thickness (mm)** | | | |
| 0.5 | 1 | 0 | 2 |
| 0.533 | 1 | 0 | 1 |
| 0.6 | 1 | 0 | 2 |
| 0.625 | 2 | 0 | 5 |
| 0.75 | 65 | 13 | 58 |
| 0.8 | 12 | 0 | 12 |
| 1 | 6 | 0 | 8 |
| 1.25 | 9 | 0 | 0 |
| 2 | 14 | 0 | 1 |
| 2.5 | 0 | 0 | 1 |
| **Manufacturer** | | | |
| Siemens | 90 | 13 | 73 |
| GE Medical Systems | 12 | 0 | 7 |
| Philips | 7 | 0 | 6 |
| Canon Medical Systems | 1 | 0 | 2 |
| Toshiba | 1 | 0 | 2 |
| **kVp (kV)** | | | |
| 70 | 21 | 3 | 21 |
| 80 | 49 | 6 | 25 |
| 90 | 17 | 4 | 10 |
| 100 | 9 | 0 | 15 |
| 110 | 2 | 0 | 5 |
| 120 | 13 | 0 | 14 |

kVp = kilovoltage peak. kV = kilovoltage.

**Supplementary Table 4: Fat attenuation index parameters in culprit versus non-culprit carotid arteries – comparing patients with versus without artifacts**

| A | Culprit lesion | | | Non-culprit lesion | | |
|---|---|---|---|---|---|---|
| FAI parameter | **No Artifacts (ICA+CCA)** | **Artifacts (ICA+CCA)** | p-value | **No Artifacts (ICA+CCA)** | **Artifacts (ICA+CCA)** | p-value |
| **Paired analysis** | n = 55 | n = 52 | | n = 55 | n = 52 | |
| FAI-PVAT unadj. (HU) | -67.4 (-74.5, -59) | -74.6 (-81.4, -67.4) | **0.01** | -68.6 (-74.6, -61.9) | -77.3 (-83.3, -66.1) | **0.002** |
| VPCI (%) | 29.8 (15, 44.2) | 20.4 (13.9, 29.1) | 0.06 | 29 (18.4, 34.9) | 20.8 (10.8, 32.5) | 0.055 |
| FAI-slope (coeff.) | -1.6 (-2.1, -1) | -1.2 (-1.6, -0.8) | **0.03** | -1.6 (-1.9, -1) | -1.1 (-1.7, -0.8) | **0.03** |
| FAI-hCCA (%) | -3.1 (-13, 10.7) | -15.7 (-26.3, 1.3) | **0.01** | -6.4 (-17.7, 5.1) | -11.6 (-20.3, -5.3) | 0.054 |
| **Unpaired analysis** | n = 56 | n = 55 | | n = 8 | n = 5 | |
| FAI-PVAT (HU) | -66.4 (-74, -60.1) | -73.5 (-79.9, -65.4) | **0.01** | -58.9 (-68.1, -57.7) | -68.1 (-69.4, -65.8) | 0.09 |
| VPCI (%) | 30.7 (15.1, 44) | 20.7 (14.2, 29.1) | 0.06 | 35.1 (28.8, 51.6) | 26.6 (26, 36.1) | 0.12 |
| FAI-slope (coeff.) | -1.6 (-2.1, -1) | -1.2 (-1.6, -0.8) | **0.04** | -2.3 (-2.8, -1.8) | -1.7 (-1.9, -1.1) | **0.04** |
| FAI-hCCA (%) | -3.9 (-12.9, 9.6) | -14.3 (-25.5, 1.6) | **0.01** | -13.9 (-22.9, -0.8) | -7.9 (-19.6, -0.1) | 0.89 |

| B | Culprit lesion | | | Non-culprit lesion | | |
|---|---|---|---|---|---|---|
| FAI parameter | **No Artifacts (ICA)** | **Artifacts (ICA)** | p-value | **No Artifacts (ICA)** | **Artifacts (ICA)** | p-value |
| **Paired analysis** | n = 55 | n = 52 | | n = 55 | n = 52 | |
| FAI-PVAT unadj. (HU) | -64.9 (-71.7, -55.2) | -75.7 (-81.2, -65) | **0.001** | -66.5 (-73.6, -58) | -73.5 (-81.4, -64.6) | **0.008** |
| VPCI (%) | 35.3 (11.1, 56.7) | 19 (11.1, 33) | **0.02** | 28.6 (16.3, 42.4) | 25.2 (12.3, 39) | 0.40 |
| FAI-slope (coeff.) | -1.7 (-2.3, -0.9) | -1.2 (-1.7, -0.8) | **0.03** | -1.4 (-1.9, -1) | -1.2 (-1.9, -0.9) | 0.62 |
| FAI-hCCA (%) | 2.7 (-11.7, 16.7) | -13.3 (-28.1, 6.4) | **0.002** | -2.4 (-18.1, 11.4) | -10.2 (-20.1, 0.2) | 0.08 |
| **Unpaired analysis** | n = 56 | n = 55 | | n = 8 | n = 5 | |
| FAI-PVAT (HU) | -61.9 (-70, -54.7) | -73.8 (-81.2, -63) | **0.001** | -61 (-62.8, -56.3) | -71.4 (-73.1, -66.2) | 0.07 |
| VPCI (%) | 36.8 (11.5, 56.4) | 18.9 (11.7, 33.1) | **0.02** | 44.3 (32.1, 59.7) | 23.7 (10.4, 35.3) | 0.12 |
| FAI-slope (coeff.) | -1.7 (-2.3, -0.9) | -1.2 (-1.7, -0.8) | **0.03** | -2.3 (-2.7, -2) | -1.5 (-1.9, -0.5) | **0.03** |
| FAI-hCCA (%) | 2.3 (-11.7, 16.6) | -12.5 (-27, 6.6) | **0.002** | 3.9 (-19.6, 9.4) | -14.1 (-21.9, 1.3) | 0.42 |

The analysis is presented for the combined segment of the internal and common carotid artery segment (A) and separately for the internal carotid artery segment (B). The results are shown as: median (first quartile, third quartile). The data was compared using two-sided t-tests.

ICA = internal carotid artery. CCA = common carotid artery. FAI = fat attenuation index. PVAT = perivascular adipose tissue. VPCI = volumetric perivascular characterization index. hCCA = healthy common carotid artery. HU = Hounsfield units. unadj. = unadjusted.

### Supplementary Figures

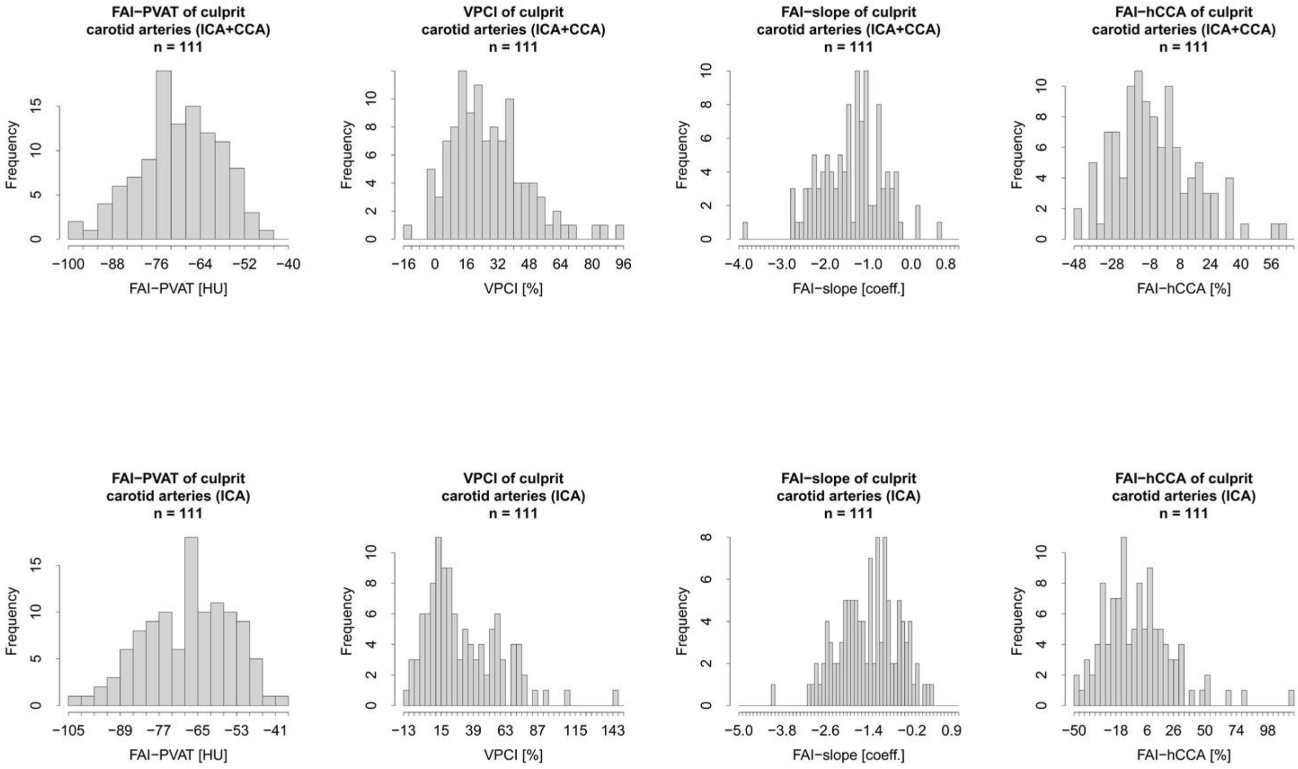


**Supplementary Figure 1: Histograms of the fat attenuation index parameters of culprit carotid arteries**

FAI = fat attenuation index. PVAT = perivascular adipose tissue. VPCI = volumetric perivascular characterization index. hCCA = healthy common carotid artery. HU = Hounsfield units. ICA = internal carotid artery. CCA = common carotid artery.

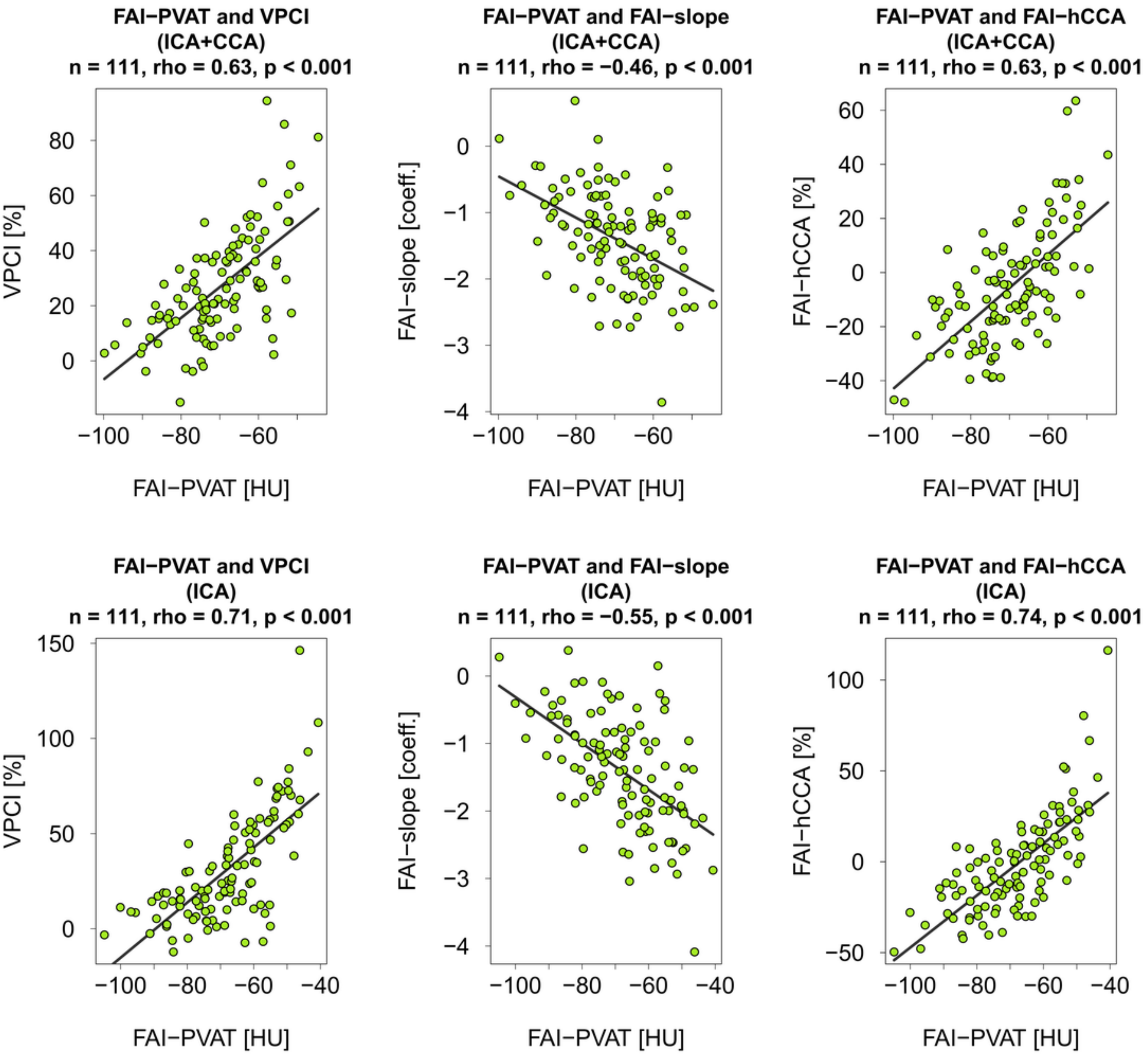


**Supplementary Figure 2: Correlation analysis between FAI-PVAT and relative PVAT metrics of culprit carotid arteries**

Spearman’s rank correlation was utilized for this analysis.

FAI = fat attenuation index. PVAT = perivascular adipose tissue. VPCI = volumetric perivascular characterization index. hCCA = healthy common carotid artery. HU = Hounsfield units. ICA = internal carotid artery. CCA = common carotid artery.

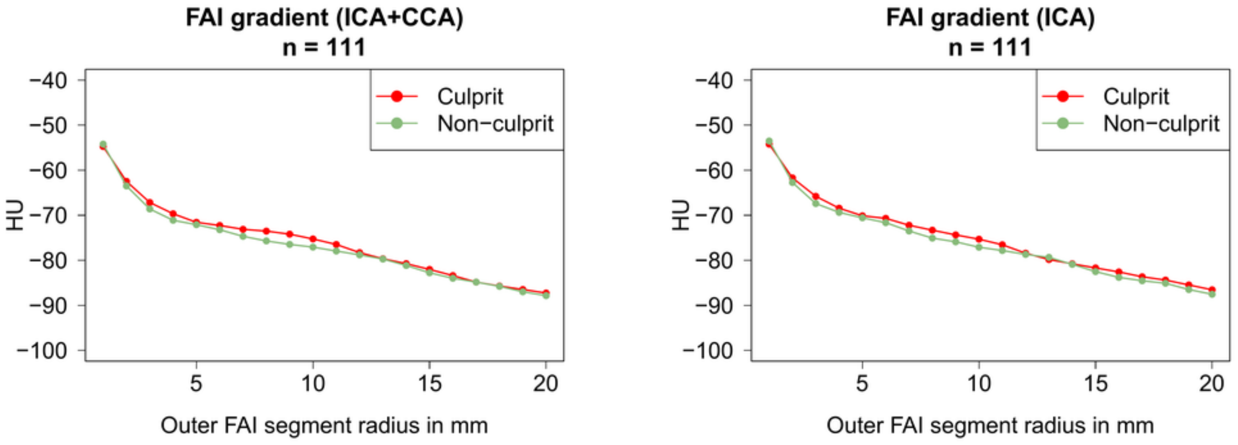


**Supplementary Figure 3: Fat attenuation index gradient of culprit carotid arteries**

For this analysis the perivascular adipose tissue at a distance of 0 to 20 mm from the arterial wall was divided into 1 mm thick adipose tissue rings. Every data point represents the FAI of one adipose tissue ring.

ICA = internal carotid artery. CCA = common carotid artery. HU = Hounsfield units.

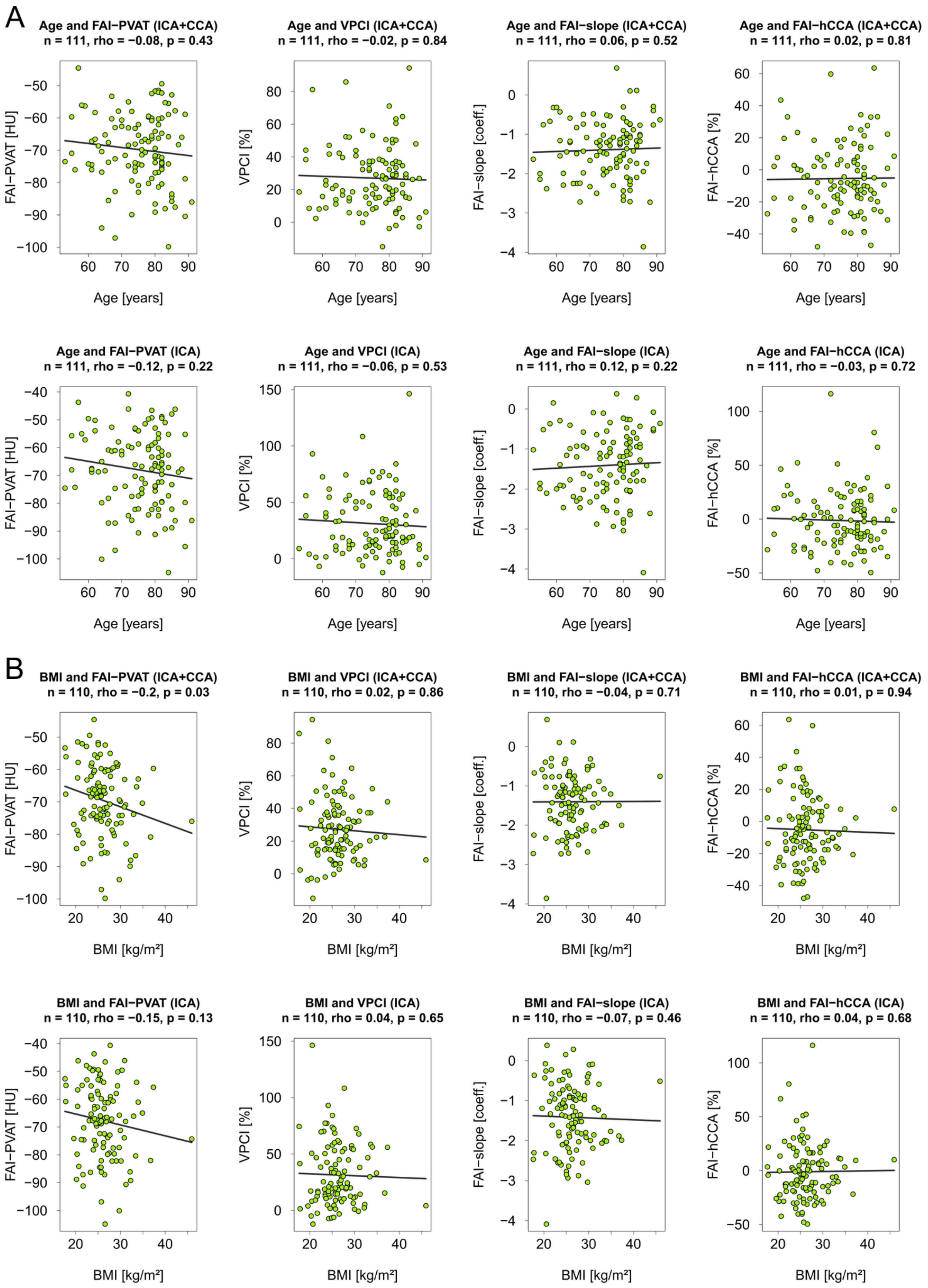

A
Age and FAI−PVAT (ICA+CCA)
n = 111, rho = −0.08, p = 0.43
Age and VPCI (ICA+CCA)
n = 111, rho = −0.02, p = 0.84
Age and FAI−slope (ICA+CCA)
n = 111, rho = 0.06, p = 0.52
Age and FAI−hCCA (ICA+CCA)
n = 111, rho = 0.02, p = 0.81
Age and FAI−PVAT (ICA)
n = 111, rho = −0.12, p = 0.22
Age and VPCI (ICA)
n = 111, rho = −0.06, p = 0.53
Age and FAI−slope (ICA)
n = 111, rho = 0.12, p = 0.22
Age and FAI−hCCA (ICA)
n = 111, rho = −0.03, p = 0.72
Age [years]
FAI−PVAT [HU]
VPCI [%]
FAI−slope [coeff.]
FAI−hCCA [%]
B
BMI and FAI−PVAT (ICA+CCA)
n = 110, rho = −0.2, p = 0.03
BMI and VPCI (ICA+CCA)
n = 110, rho = 0.02, p = 0.86
BMI and FAI−slope (ICA+CCA)
n = 110, rho = −0.04, p = 0.71
BMI and FAI−hCCA (ICA+CCA)
n = 110, rho = 0.01, p = 0.94
BMI and FAI−PVAT (ICA)
n = 110, rho = −0.15, p = 0.13
BMI and VPCI (ICA)
n = 110, rho = 0.04, p = 0.65
BMI and FAI−slope (ICA)
n = 110, rho = −0.07, p = 0.46
BMI and FAI−hCCA (ICA)
n = 110, rho = 0.04, p = 0.68
BMI [kg/m²]

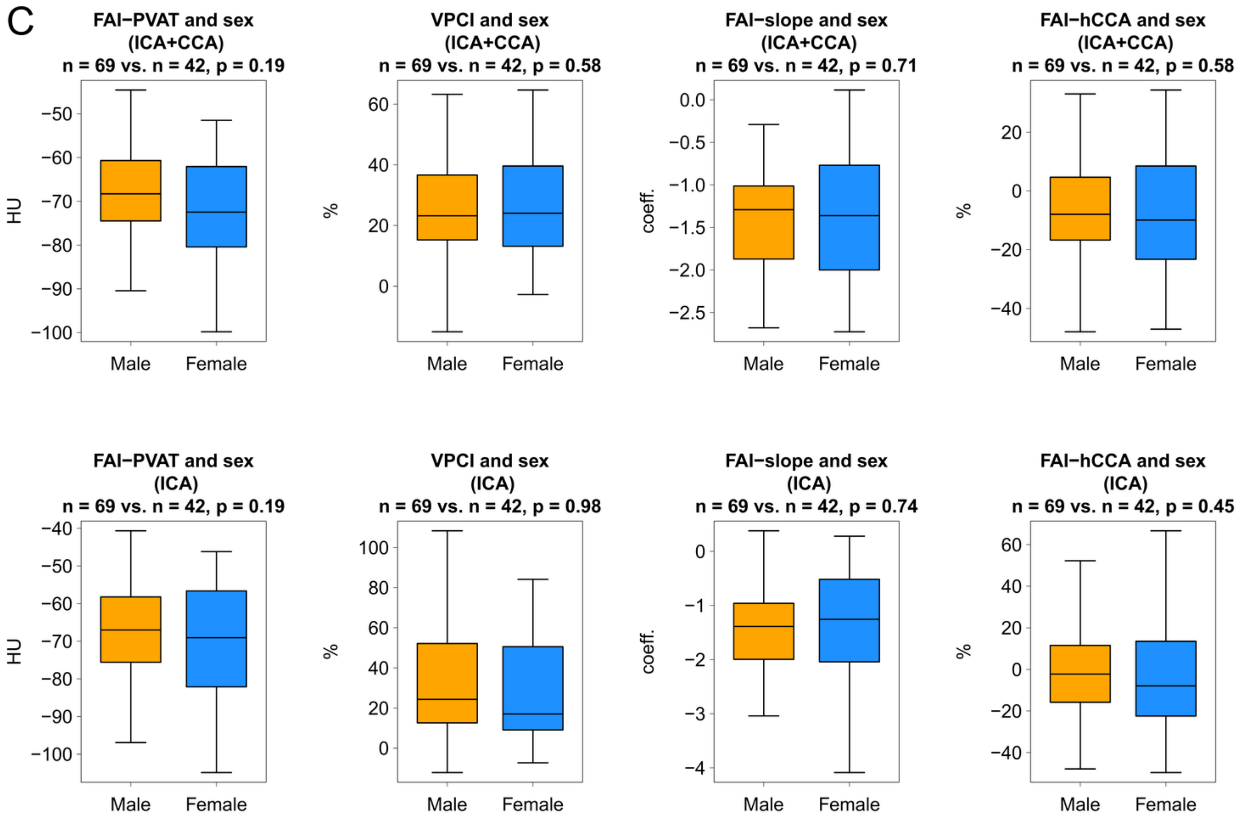


**Supplementary Figure 4: Relationship of fat attenuation index parameters and baseline characteristics in culprit carotid arteries**

Shown is the relation of Fat attenuation index (FAI) parameters and age (A), BMI (B) and sex (C). Every analysis is presented for the internal and common carotid artery segment in the upper row and separately for the internal carotid artery in the lower row. Spearman's rank correlation was utilized for age and BMI and two-sided t-tests for sex.

PVAT = perivascular adipose tissue. VPCI = volumetric perivascular characterization index. hCCA = healthy common carotid artery. HU = Hounsfield units. ICA = internal carotid artery. CCA = common carotid artery.

A

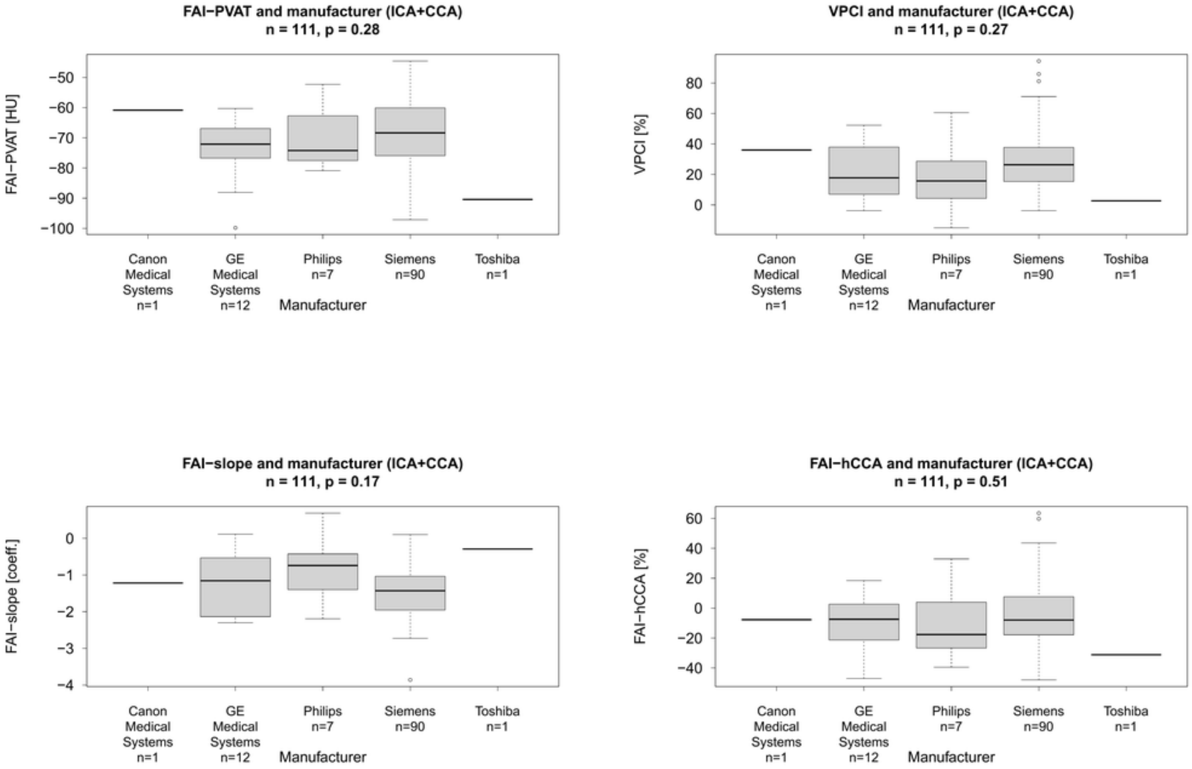
FAI−PVAT and manufacturer (ICA+CCA)
n = 111, p = 0.28
FAI−PVAT [HU]
VPCI and manufacturer (ICA+CCA)
n = 111, p = 0.27
VPCI [%]
FAI−slope and manufacturer (ICA+CCA)
n = 111, p = 0.17
FAI−slope [coeff.]
FAI−hCCA and manufacturer (ICA+CCA)
n = 111, p = 0.51
FAI−hCCA [%]
Canon Medical Systems n=1
GE Medical Systems n=12
Philips n=7
Siemens n=90
Toshiba n=1
Manufacturer

B

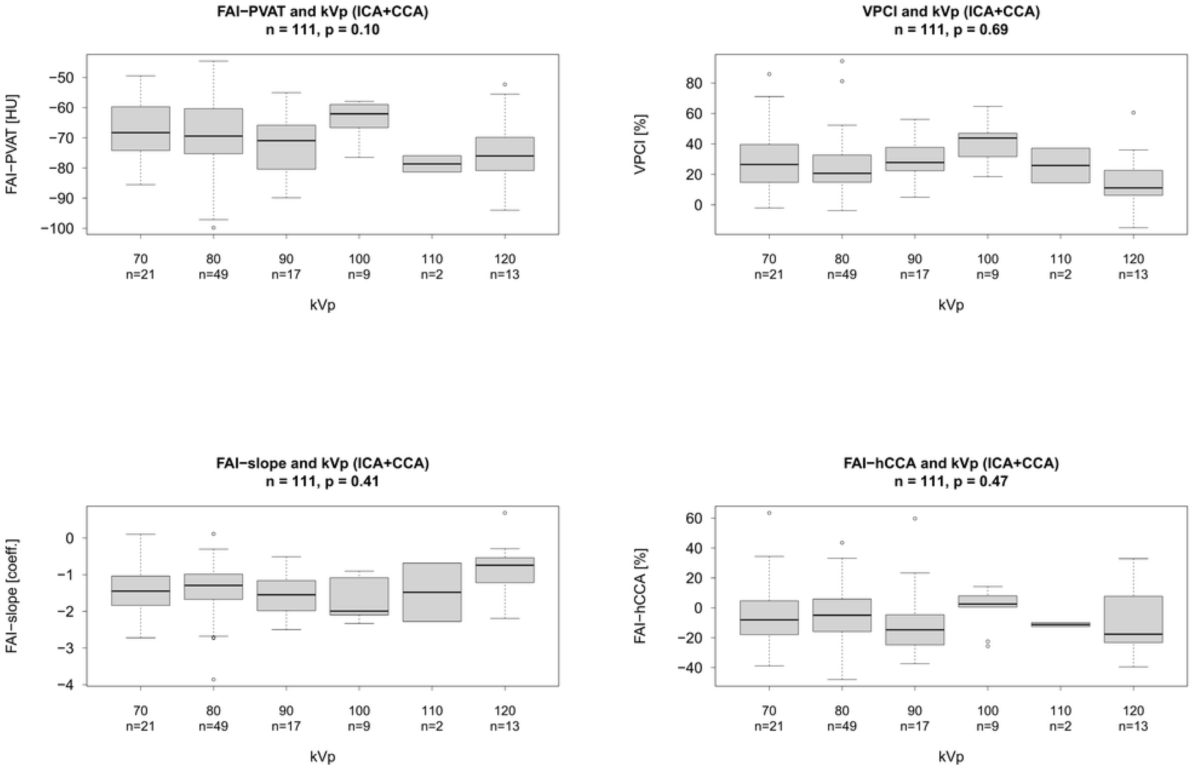
FAI−PVAT and kVp (ICA+CCA)
n = 111, p = 0.10
FAI−PVAT [HU]
VPCI and kVp (ICA+CCA)
n = 111, p = 0.69
VPCI [%]
FAI−slope and kVp (ICA+CCA)
n = 111, p = 0.41
FAI−slope [coeff.]
FAI−hCCA and kVp (ICA+CCA)
n = 111, p = 0.47
FAI−hCCA [%]
70 n=21
80 n=49
90 n=17
100 n=9
110 n=2
120 n=13
kVp

C

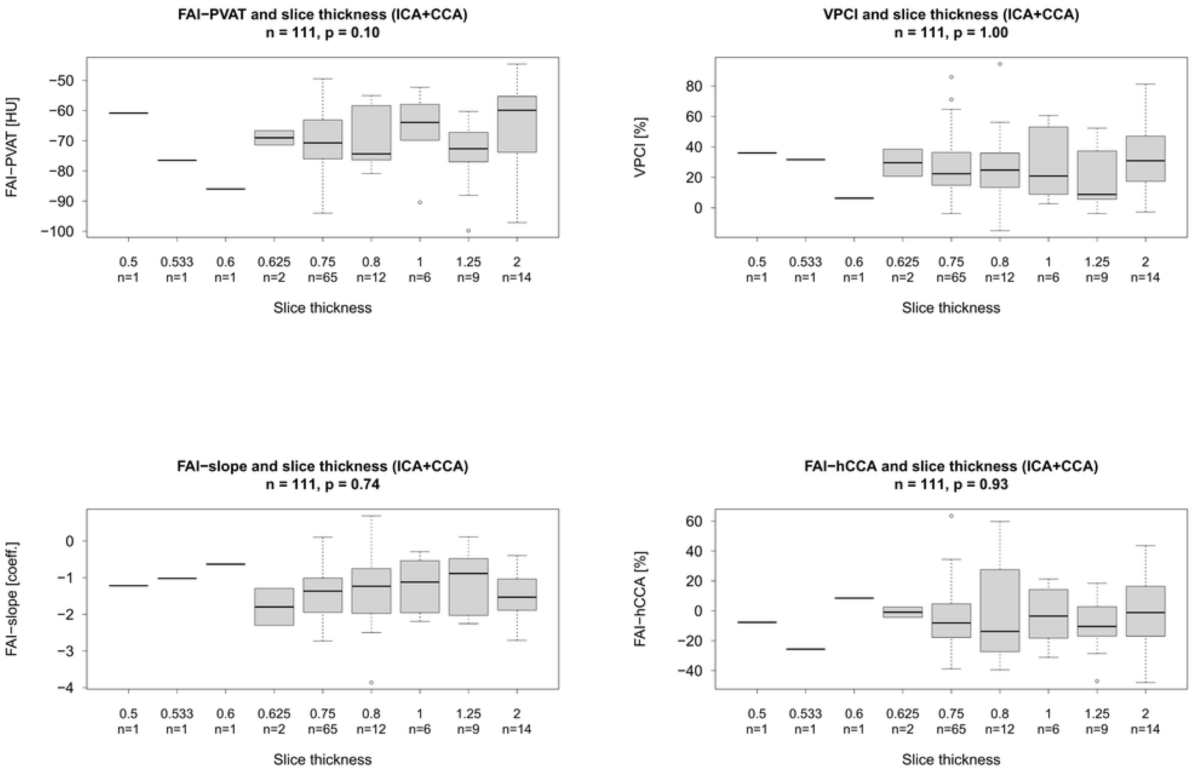


**Supplementary Figure 5: Relation of fat attenuation index parameters and CT characteristics in culprit carotid arteries**

Comparison of fat attenuation index (FAI) parameters and CT manufacturer (A), kilovoltage peak (B) and CT slice thickness (C). The Kruskal-Wallis test was used for the analysis of FAI in relation to the manufacturer and the Jonckheere-Terpstra test for kilovoltage peak and slice thickness.

PVAT = perivascular adipose tissue. VPCI = volumetric perivascular characterization index. hCCA = healthy common carotid artery. HU = Hounsfield units. ICA = internal carotid artery. CCA = common carotid artery. kVp = kilovoltage peak.

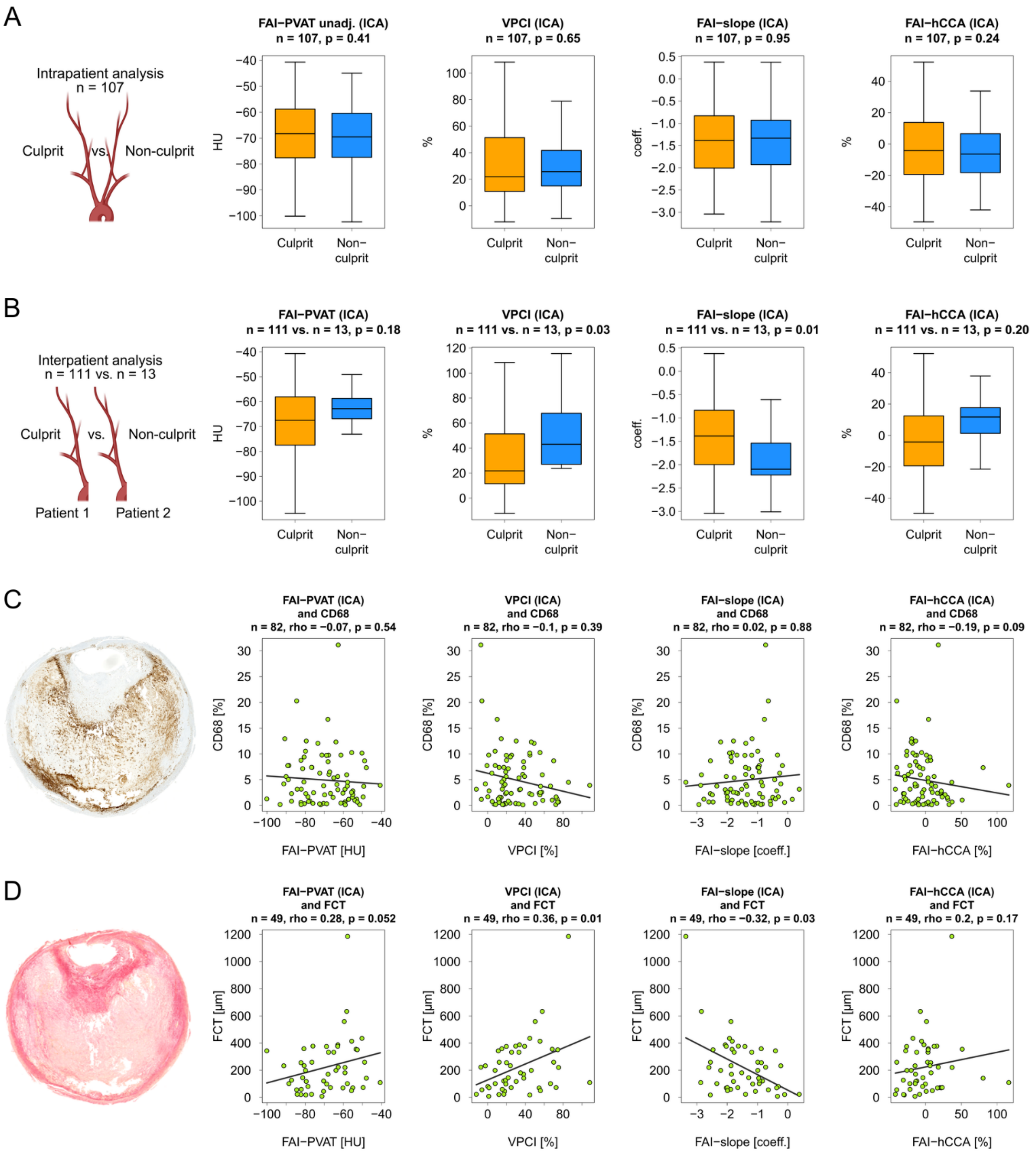
A
Intrapatient analysis
n = 107
Culprit
vs.
Non-culprit
FAI−PVAT unadj. (ICA)
n = 107, p = 0.41
VPCI (ICA)
n = 107, p = 0.65
FAI−slope (ICA)
n = 107, p = 0.95
FAI−hCCA (ICA)
n = 107, p = 0.24
HU
%
coeff.
Culprit
Non−culprit
B
Interpatient analysis
n = 111 vs. n = 13
Patient 1
Patient 2
FAI−PVAT (ICA)
n = 111 vs. n = 13, p = 0.18
VPCI (ICA)
n = 111 vs. n = 13, p = 0.03
FAI−slope (ICA)
n = 111 vs. n = 13, p = 0.01
FAI−hCCA (ICA)
n = 111 vs. n = 13, p = 0.20
C
FAI−PVAT (ICA) and CD68
n = 82, rho = −0.07, p = 0.54
VPCI (ICA) and CD68
n = 82, rho = −0.1, p = 0.39
FAI−slope (ICA) and CD68
n = 82, rho = 0.02, p = 0.88
FAI−hCCA (ICA) and CD68
n = 82, rho = −0.19, p = 0.09
CD68 [%]
FAI−PVAT [HU]
VPCI [%]
FAI−slope [coeff.]
FAI−hCCA [%]
D
FAI−PVAT (ICA) and FCT
n = 49, rho = 0.28, p = 0.052
VPCI (ICA) and FCT
n = 49, rho = 0.36, p = 0.01
FAI−slope (ICA) and FCT
n = 49, rho = −0.32, p = 0.03
FAI−hCCA (ICA) and FCT
n = 49, rho = 0.2, p = 0.17
FCT [µm]

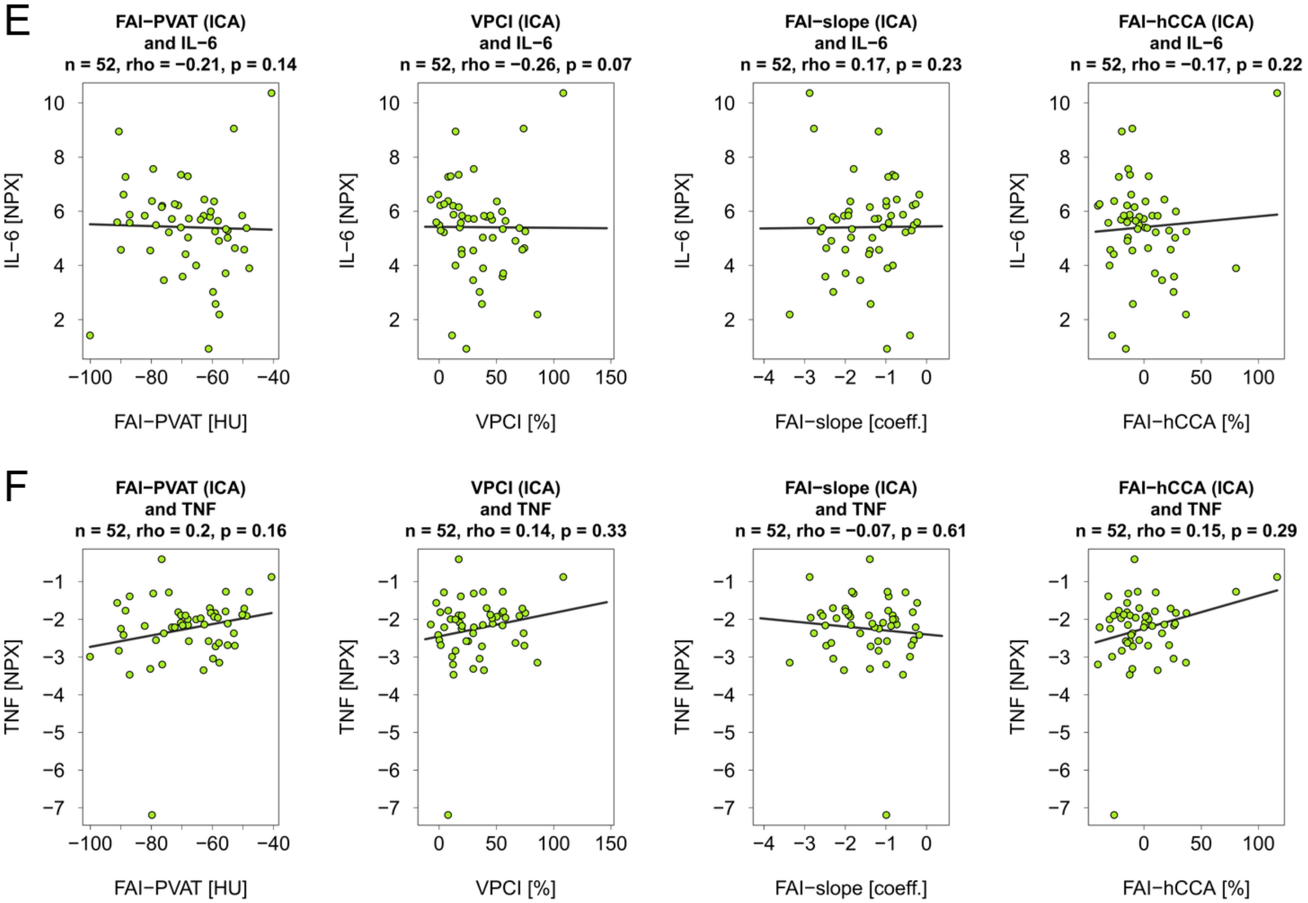


**Supplementary Figure 6: Fat attenuation index parameters in the internal carotid artery**

Analysis of the fat attenuation index (FAI) parameters only around a 2 cm segment of the internal carotid artery (ICA), leaving out the common carotid artery (CCA). Paired analysis of FAI parameters in culprit versus non-culprit carotid arteries (A). Unpaired analysis of culprit carotid arteries versus a random non-culprit side of non-carotid stroke patients (B). Relationship of FAI parameters and CD68 (C), fibrous cap thickness (D), IL-6 (E) and TNF (F), analyzed with Spearman's rank correlation.

FAI = fat attenuation index. PVAT = perivascular adipose tissue. VPCI = volumetric perivascular characterization index. hCCA = healthy common carotid artery. HU = Hounsfield units. unadj. = unadjusted. FCT = fibrous cap thickness. IL-6 = interleukin 6. TNF = Tumor necrosis factor. NPX = normalized protein expression.

Created in part with BioRender. Bernhagen, L. (2026) https://BioRender.com/gchun17 and https://BioRender.com/yi80xa4

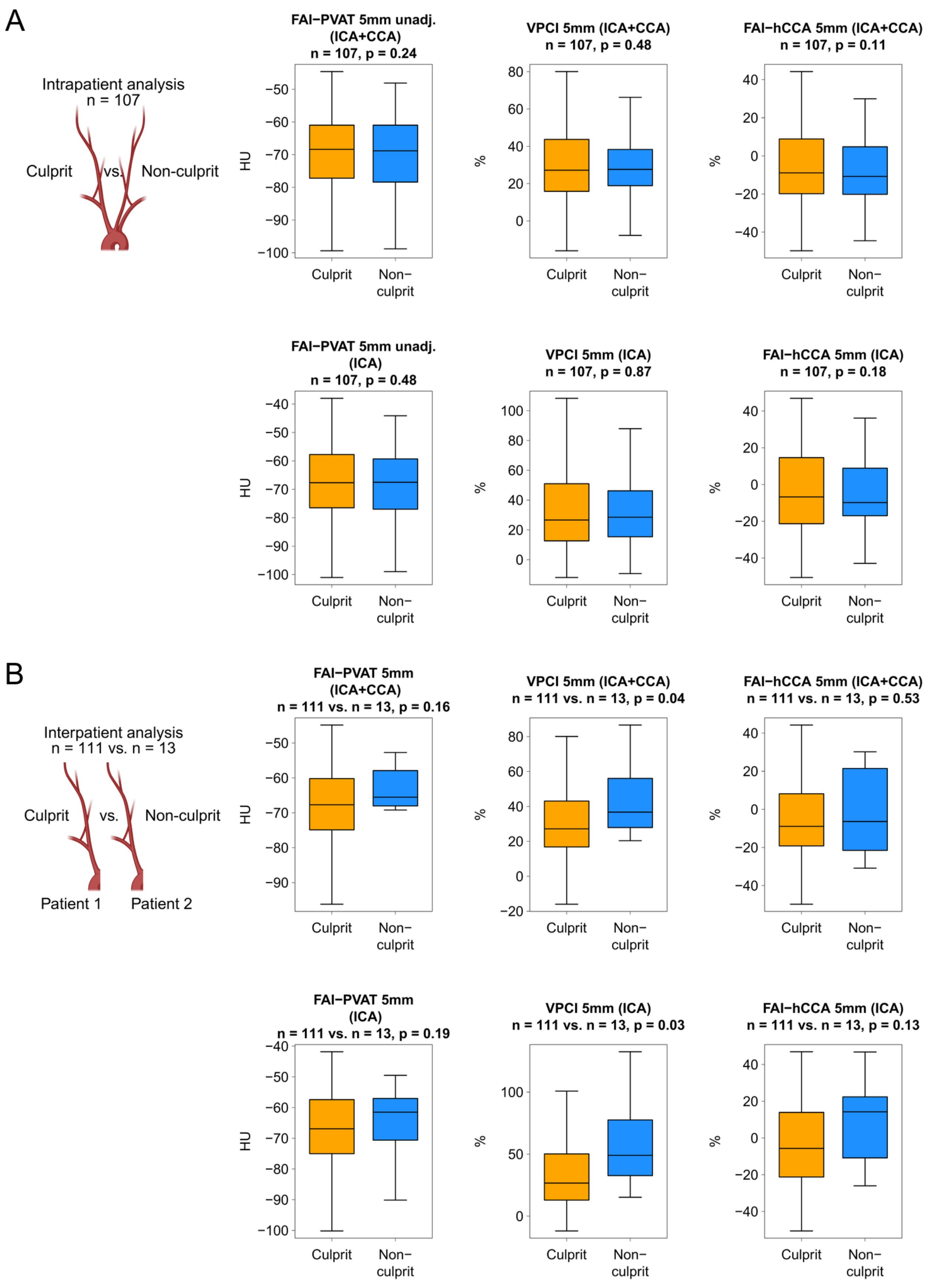

A
Intrapatient analysis
n = 107
Culprit
vs.
Non-culprit
FAI−PVAT 5mm unadj. (ICA+CCA)
n = 107, p = 0.24
VPCI 5mm (ICA+CCA)
n = 107, p = 0.48
FAI−hCCA 5mm (ICA+CCA)
n = 107, p = 0.11
FAI−PVAT 5mm unadj. (ICA)
n = 107, p = 0.48
VPCI 5mm (ICA)
n = 107, p = 0.87
FAI−hCCA 5mm (ICA)
n = 107, p = 0.18
B
Interpatient analysis
n = 111 vs. n = 13
Culprit
vs.
Non-culprit
Patient 1
Patient 2
FAI−PVAT 5mm (ICA+CCA)
n = 111 vs. n = 13, p = 0.16
VPCI 5mm (ICA+CCA)
n = 111 vs. n = 13, p = 0.04
FAI−hCCA 5mm (ICA+CCA)
n = 111 vs. n = 13, p = 0.53
FAI−PVAT 5mm (ICA)
n = 111 vs. n = 13, p = 0.19
VPCI 5mm (ICA)
n = 111 vs. n = 13, p = 0.03
FAI−hCCA 5mm (ICA)
n = 111 vs. n = 13, p = 0.13
HU
%
Culprit
Non−culprit

C

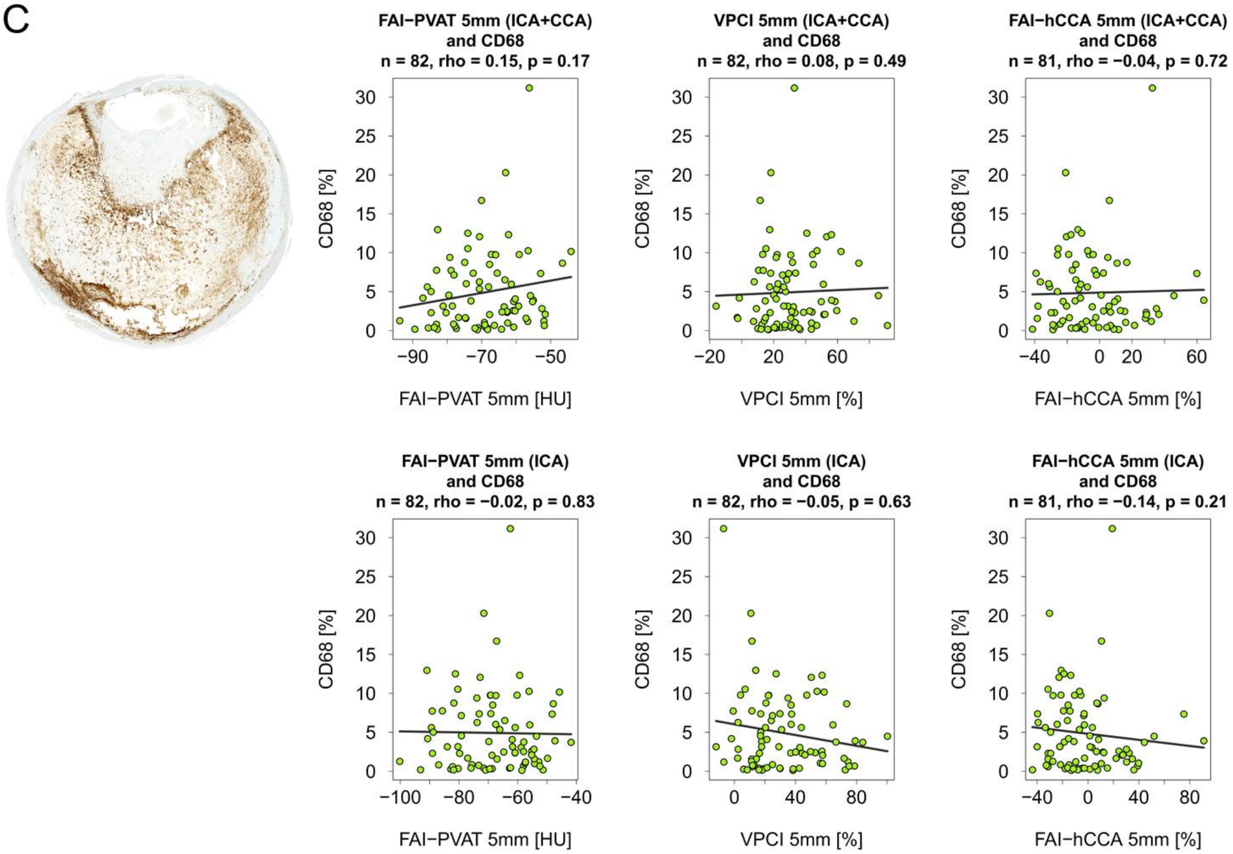
FAI−PVAT 5mm (ICA+CCA)
and CD68
n = 82, rho = 0.15, p = 0.17
VPCI 5mm (ICA+CCA)
and CD68
n = 82, rho = 0.08, p = 0.49
FAI−hCCA 5mm (ICA+CCA)
and CD68
n = 81, rho = −0.04, p = 0.72
FAI−PVAT 5mm (ICA)
and CD68
n = 82, rho = −0.02, p = 0.83
VPCI 5mm (ICA)
and CD68
n = 82, rho = −0.05, p = 0.63
FAI−hCCA 5mm (ICA)
and CD68
n = 81, rho = −0.14, p = 0.21
CD68 [%]
FAI−PVAT 5mm [HU]
VPCI 5mm [%]
FAI−hCCA 5mm [%]

D

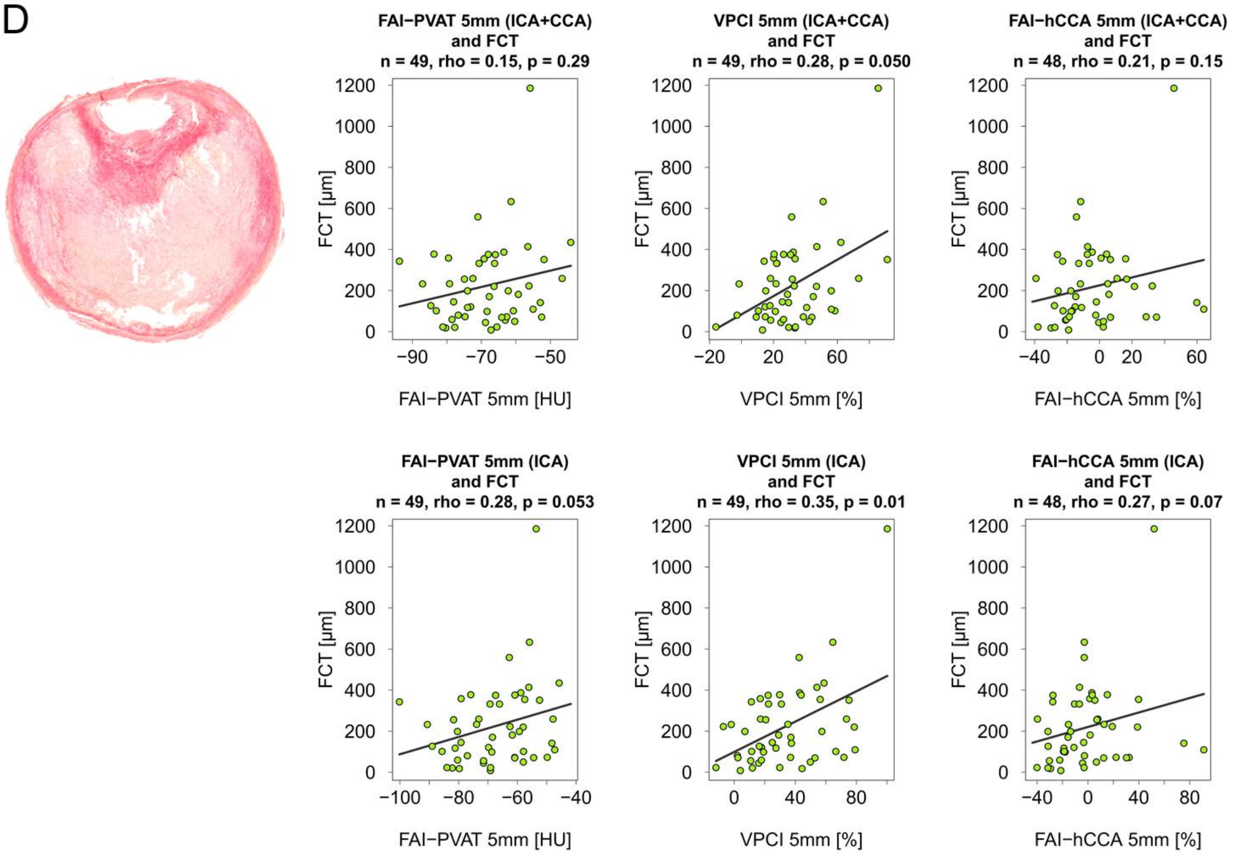
FAI−PVAT 5mm (ICA+CCA)
and FCT
n = 49, rho = 0.15, p = 0.29
VPCI 5mm (ICA+CCA)
and FCT
n = 49, rho = 0.28, p = 0.050
FAI−hCCA 5mm (ICA+CCA)
and FCT
n = 48, rho = 0.21, p = 0.15
FAI−PVAT 5mm (ICA)
and FCT
n = 49, rho = 0.28, p = 0.053
VPCI 5mm (ICA)
and FCT
n = 49, rho = 0.35, p = 0.01
FAI−hCCA 5mm (ICA)
and FCT
n = 48, rho = 0.27, p = 0.07
FCT [µm]
FAI−PVAT 5mm [HU]
VPCI 5mm [%]
FAI−hCCA 5mm [%]

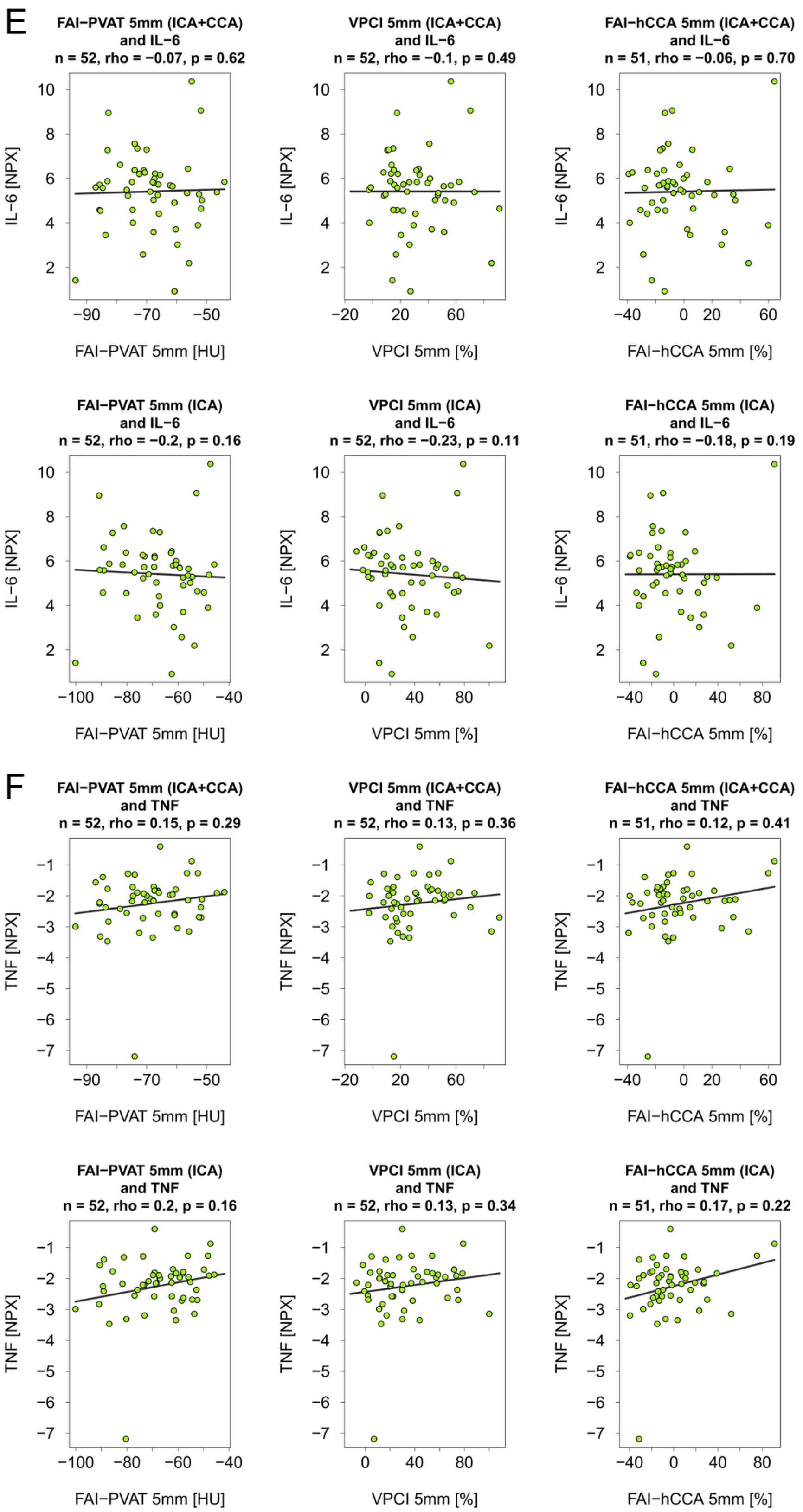
E
FAI−PVAT 5mm (ICA+CCA) and IL−6
n = 52, rho = −0.07, p = 0.62
VPCI 5mm (ICA+CCA) and IL−6
n = 52, rho = −0.1, p = 0.49
FAI−hCCA 5mm (ICA+CCA) and IL−6
n = 51, rho = −0.06, p = 0.70
FAI−PVAT 5mm (ICA) and IL−6
n = 52, rho = −0.2, p = 0.16
VPCI 5mm (ICA) and IL−6
n = 52, rho = −0.23, p = 0.11
FAI−hCCA 5mm (ICA) and IL−6
n = 51, rho = −0.18, p = 0.19
IL−6 [NPX]
FAI−PVAT 5mm [HU]
VPCI 5mm [%]
FAI−hCCA 5mm [%]
F
FAI−PVAT 5mm (ICA+CCA) and TNF
n = 52, rho = 0.15, p = 0.29
VPCI 5mm (ICA+CCA) and TNF
n = 52, rho = 0.13, p = 0.36
FAI−hCCA 5mm (ICA+CCA) and TNF
n = 51, rho = 0.12, p = 0.41
FAI−PVAT 5mm (ICA) and TNF
n = 52, rho = 0.2, p = 0.16
VPCI 5mm (ICA) and TNF
n = 52, rho = 0.13, p = 0.34
FAI−hCCA 5mm (ICA) and TNF
n = 51, rho = 0.17, p = 0.22
TNF [NPX]

**Supplementary Figure 7: Fat attenuation index parameters with a 5 mm radius of perivascular adipose tissue**

Alternative definition of the perivascular adipose tissue using a fixed radius of 5 mm measured from the outer vessel edge, instead of incorporating the mean diameter of the vessel segment, which is then utilized to calculate FAI-PVAT 5mm, VPCI 5mm and FAI-hCCA 5mm. Paired analysis of FAI parameters comparing culprit and non-culprit carotid arteries (A). Unpaired analysis showing the comparison of culprit carotid arteries versus a random non-culprit side of non-carotid stroke patients (B). Analysis of the relationship of FAI parameters and CD68 (C), fibrous cap thickness (D), IL-6 (E) and TNF (F), utilizing Spearman's rank correlation. For each analysis, the results are shown for the combined segment of the internal and common carotid artery in the upper row and separately for the internal carotid artery in the lower row. One asymptomatic patient was excluded from the FAI-hCCA 5mm correlation analyses due to not having any adipose tissue in a radius of 5 mm around the hCCA-Segment.

FAI = fat attenuation index. PVAT = perivascular adipose tissue. VPCI = volumetric perivascular characterization index. hCCA = healthy common carotid artery. HU = Hounsfield units. unadj. = unadjusted. FCT = fibrous cap thickness. IL-6 = interleukin 6. TNF = Tumor necrosis factor. NPX = normalized protein expression.

Created in part with BioRender. Bernhagen, L. (2026) https://BioRender.com/gchun17 and https://BioRender.com/yi80xa4

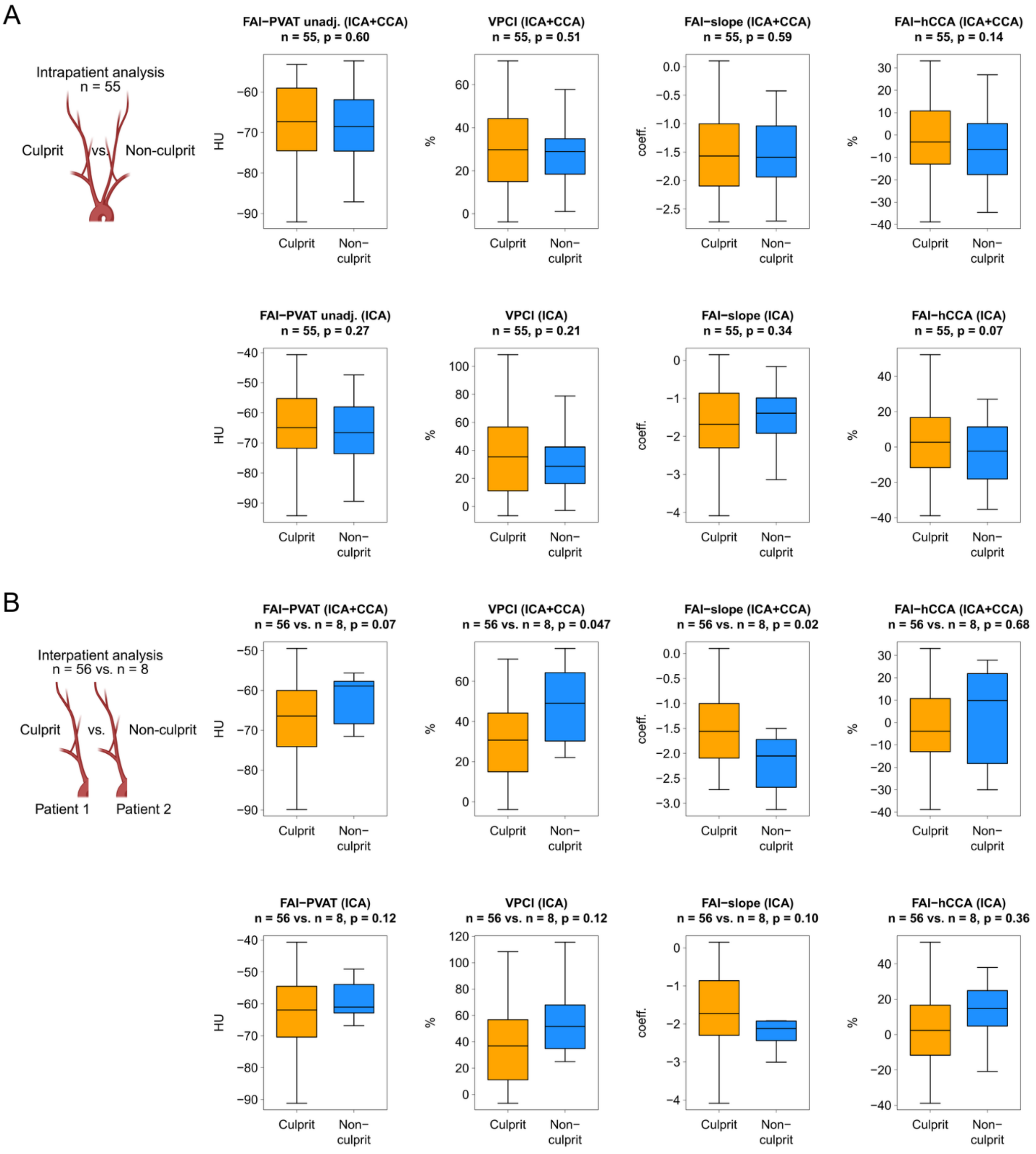
A
Intrapatient analysis
n = 55
Culprit
vs.
Non-culprit
FAI−PVAT unadj. (ICA+CCA)
n = 55, p = 0.60
VPCI (ICA+CCA)
n = 55, p = 0.51
FAI−slope (ICA+CCA)
n = 55, p = 0.59
FAI−hCCA (ICA+CCA)
n = 55, p = 0.14
FAI−PVAT unadj. (ICA)
n = 55, p = 0.27
VPCI (ICA)
n = 55, p = 0.21
FAI−slope (ICA)
n = 55, p = 0.34
FAI−hCCA (ICA)
n = 55, p = 0.07
B
Interpatient analysis
n = 56 vs. n = 8
Culprit
vs.
Non-culprit
Patient 1
Patient 2
FAI−PVAT (ICA+CCA)
n = 56 vs. n = 8, p = 0.07
VPCI (ICA+CCA)
n = 56 vs. n = 8, p = 0.047
FAI−slope (ICA+CCA)
n = 56 vs. n = 8, p = 0.02
FAI−hCCA (ICA+CCA)
n = 56 vs. n = 8, p = 0.68
FAI−PVAT (ICA)
n = 56 vs. n = 8, p = 0.12
VPCI (ICA)
n = 56 vs. n = 8, p = 0.12
FAI−slope (ICA)
n = 56 vs. n = 8, p = 0.10
FAI−hCCA (ICA)
n = 56 vs. n = 8, p = 0.36
HU
%
coeff.
Culprit
Non−culprit

C

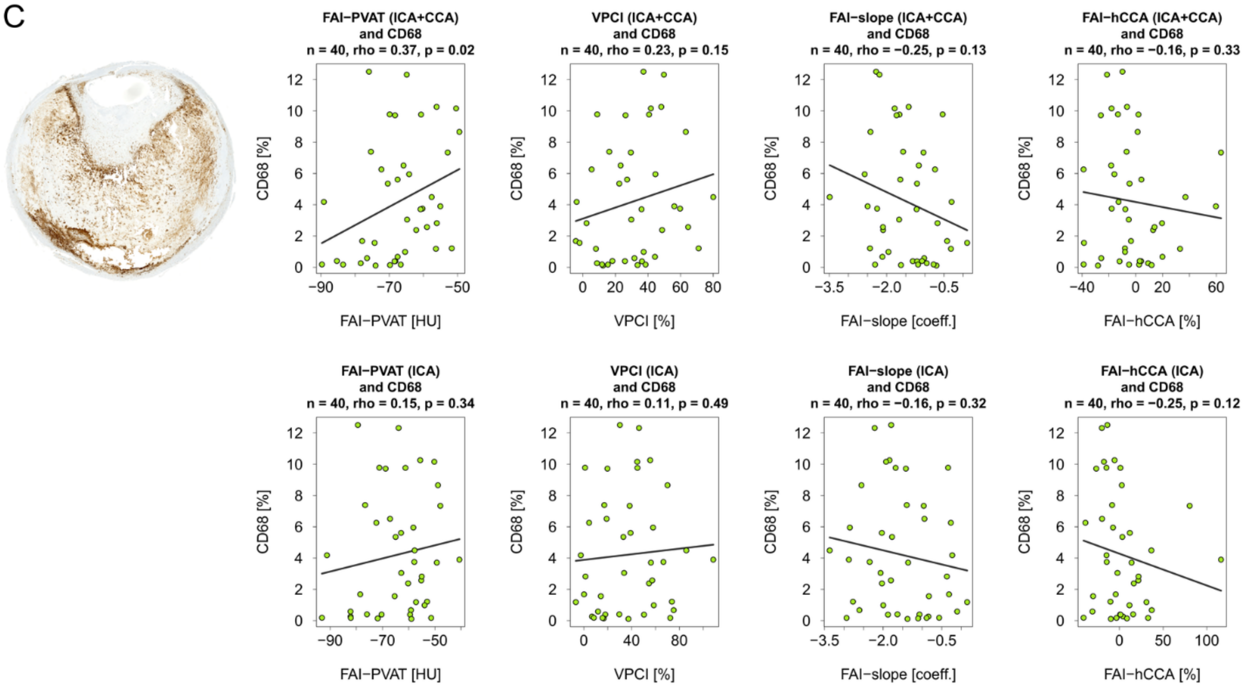
FAI−PVAT (ICA+CCA) and CD68
n = 40, rho = 0.37, p = 0.02
VPCI (ICA+CCA) and CD68
n = 40, rho = 0.23, p = 0.15
FAI−slope (ICA+CCA) and CD68
n = 40, rho = −0.25, p = 0.13
FAI−hCCA (ICA+CCA) and CD68
n = 40, rho = −0.16, p = 0.33
FAI−PVAT (ICA) and CD68
n = 40, rho = 0.15, p = 0.34
VPCI (ICA) and CD68
n = 40, rho = 0.11, p = 0.49
FAI−slope (ICA) and CD68
n = 40, rho = −0.16, p = 0.32
FAI−hCCA (ICA) and CD68
n = 40, rho = −0.25, p = 0.12
CD68 [%]
FAI−PVAT [HU]
VPCI [%]
FAI−slope [coeff.]
FAI−hCCA [%]

D

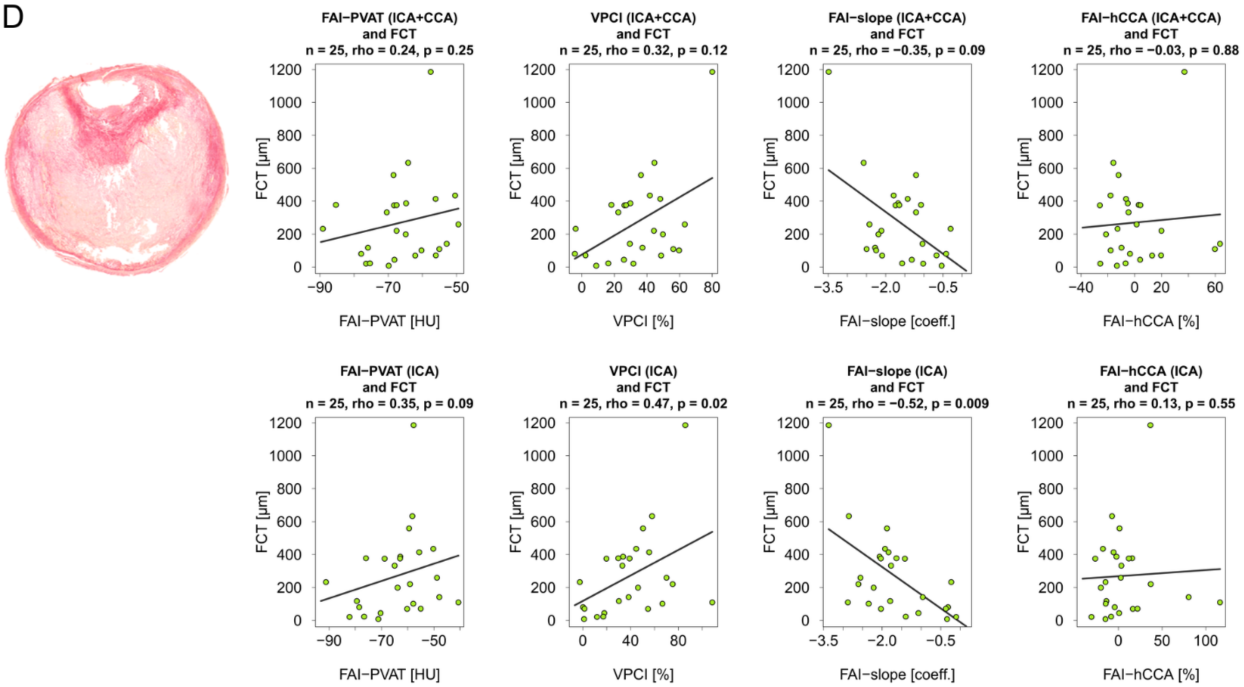
FAI−PVAT (ICA+CCA) and FCT
n = 25, rho = 0.24, p = 0.25
VPCI (ICA+CCA) and FCT
n = 25, rho = 0.32, p = 0.12
FAI−slope (ICA+CCA) and FCT
n = 25, rho = −0.35, p = 0.09
FAI−hCCA (ICA+CCA) and FCT
n = 25, rho = −0.03, p = 0.88
FAI−PVAT (ICA) and FCT
n = 25, rho = 0.35, p = 0.09
VPCI (ICA) and FCT
n = 25, rho = 0.47, p = 0.02
FAI−slope (ICA) and FCT
n = 25, rho = −0.52, p = 0.009
FAI−hCCA (ICA) and FCT
n = 25, rho = 0.13, p = 0.55
FCT [µm]
FAI−PVAT [HU]
VPCI [%]
FAI−slope [coeff.]
FAI−hCCA [%]

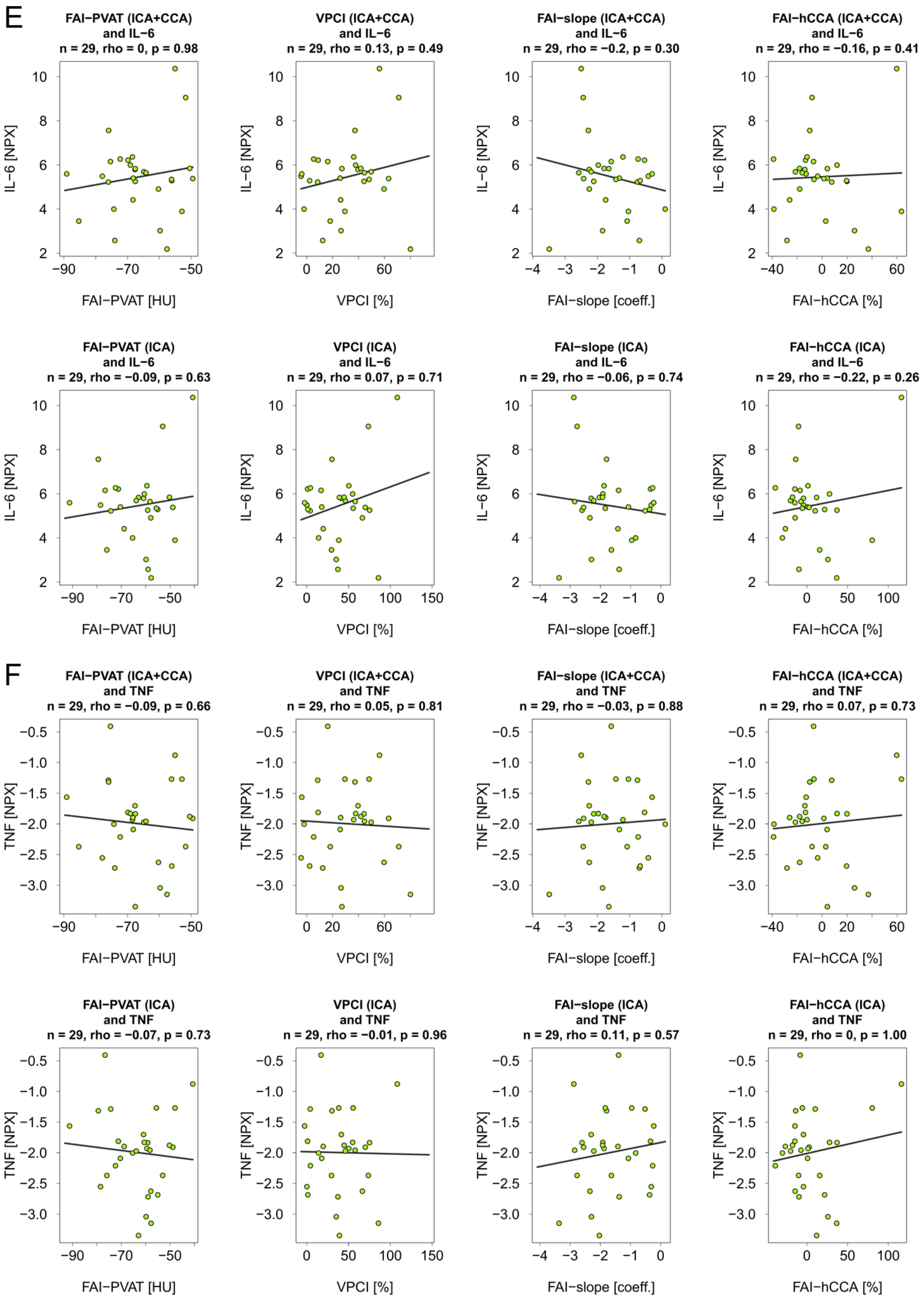
E
FAI−PVAT (ICA+CCA)
and IL−6
n = 29, rho = 0, p = 0.98
VPCI (ICA+CCA)
and IL−6
n = 29, rho = 0.13, p = 0.49
FAI−slope (ICA+CCA)
and IL−6
n = 29, rho = −0.2, p = 0.30
FAI−hCCA (ICA+CCA)
and IL−6
n = 29, rho = −0.16, p = 0.41
FAI−PVAT (ICA)
and IL−6
n = 29, rho = −0.09, p = 0.63
VPCI (ICA)
and IL−6
n = 29, rho = 0.07, p = 0.71
FAI−slope (ICA)
and IL−6
n = 29, rho = −0.06, p = 0.74
FAI−hCCA (ICA)
and IL−6
n = 29, rho = −0.22, p = 0.26
IL−6 [NPX]
FAI−PVAT [HU]
VPCI [%]
FAI−slope [coeff.]
FAI−hCCA [%]
F
FAI−PVAT (ICA+CCA)
and TNF
n = 29, rho = −0.09, p = 0.66
VPCI (ICA+CCA)
and TNF
n = 29, rho = 0.05, p = 0.81
FAI−slope (ICA+CCA)
and TNF
n = 29, rho = −0.03, p = 0.88
FAI−hCCA (ICA+CCA)
and TNF
n = 29, rho = 0.07, p = 0.73
FAI−PVAT (ICA)
and TNF
n = 29, rho = −0.07, p = 0.73
VPCI (ICA)
and TNF
n = 29, rho = −0.01, p = 0.96
FAI−slope (ICA)
and TNF
n = 29, rho = 0.11, p = 0.57
FAI−hCCA (ICA)
and TNF
n = 29, rho = 0, p = 1.00
TNF [NPX]
FAI−PVAT [HU]
VPCI [%]
FAI−slope [coeff.]
FAI−hCCA [%]

**Supplementary Figure 8: Fat attenuation index parameters of patients without CT image artifacts**

Paired sample, comparing the FAI parameters of culprit versus non-culprit carotid arteries (A). Unpaired sample, comparing the FAI of culprit arteries versus a random non-culprit side of non-carotid stroke patients (B). The relationship between FAI parameters and CD68 (C), fibrous cap thickness (D), IL-6 (E) and TNF (F) are shown. For each analysis, the results are presented for a combined segment of the internal and common carotid artery in the upper row and separately for a segment only containing the internal carotid artery in the lower row.

FAI = fat attenuation index. PVAT = perivascular adipose tissue. VPCI = volumetric perivascular characterization index. hCCA = healthy common carotid artery. HU = Hounsfield units. unadj. = unadjusted. FCT = fibrous cap thickness. IL-6 = interleukin 6. TNF = Tumor necrosis factor. NPX = normalized protein expression.

Created in part with BioRender. Bernhagen, L. (2026) https://BioRender.com/ca9ev7p and https://BioRender.com/p81j1rv
